# Neurodevelopmental, cognitive, behavioural and mental health impairments following childhood malnutrition: a systematic review

**DOI:** 10.1101/2022.04.11.22273736

**Authors:** Amir Kirolos, Magdalena Goyheneix, Mike Kalmus Eliasz, Mphatso Chisala, Samantha Lissauer, Melissa Gladstone, Marko Kerac

## Abstract

**Background:** Severe childhood malnutrition impairs growth and development short-term, but current understanding of long-term outcomes is limited. We aimed to identify studies assessing neurodevelopmental, cognitive, behavioural and mental health outcomes following childhood malnutrition.

**Methods:** We systematically searched MEDLINE, EMBASE, Global Health and PsychINFO for studies assessing these outcomes in those exposed to childhood malnutrition in low- and middle-income settings. We included studies assessing undernutrition measured by low mid-upper arm circumference, weight-for-height, weight-for-age or nutritional oedema. We used guidelines for synthesis of results without meta-analysis to analyse three outcome areas: neurodevelopment, cognition/academic achievement, behaviour/mental health.

**Results:** We identified 30 studies, including some long-term cohorts reporting outcomes through to adulthood. There is strong evidence that malnutrition in childhood negatively impacts neurodevelopment based on high-quality studies using validated neurodevelopmental assessment tools. There is also strong evidence that malnutrition impairs academic achievement with agreement across seven studies investigating this outcome. 8 of 11 studies showed association between childhood malnutrition and impaired cognition. This moderate evidence is limited by some studies failing to measure important confounders such as socioeconomic status. 5 of 7 studies found a difference in behavioural assessment scores in those exposed to childhood malnutrition compared to controls but this moderate evidence is similarly limited by unmeasured confounders. Mental health impacts were difficult to ascertain due to few studies with mixed results.

**Conclusions:** Childhood malnutrition is associated with impaired neurodevelopment, academic achievement, cognition and behavioural problems but evidence regarding possible mental health impacts is inconclusive. Future research should explore the interplay of childhood and later-life adversities on these outcomes. Whilst evidence on improving nutritional and clinical therapies to reduce long-term risks is also needed, preventing and eliminating child malnutrition is likely to be the best way of preventing long-term neurocognitive harms.

**PROSPERO registration number:** CRD42021260498

**KEY QUESTIONS:** *What is already known?:*

– High mortality risk and impaired growth are well-recognised short-term risks of childhood malnutrition
– Whilst there is increasing appreciation of longer-term risks for survivors, notably adult cardiometabolic non-communicable disease, other longer-term risks have been poorly described.

*What are the new findings?:*

– There is strong evidence that malnutrition impairs neurodevelopment and academic achievement in childhood which has significant implications for future wellbeing and prospects of those affected
– Childhood malnutrition is associated with impaired cognition and behavioural problems with evidence of effects through to adolescence and adulthood but the effect of nutritional treatment and interplay with childhood adversity, co-existing illness such as HIV and environmental factors in influencing these outcomes is unclear

*What do the new findings imply?:*

– Study findings imply that there are likely to be long-term effects of childhood malnutrition on cognition and wellbeing lasting through adolescence and adulthood
– Long-term needs of malnutrition survivors need to be carefully considered in treatment programmes. Further research is needed on the effects of nutritional therapy, adversity and environmental factors to tailor future interventions, particularly with regards to mental health which has been little researched

## INTRODUCTION

Severe childhood malnutrition is widespread and has a high disease burden concentrated in low- and middle-income settings^1^. To date, most malnutrition policies and treatment programmes have focused on short-term risks, notably infections and death^2, 3^. There is however growing evidence of long-term risks for malnutrition survivors, including that of later-life non-communicable disease ^4^. The prevalence of malnutrition has decreased in recent years due to concerted global efforts ^1^. However, climate change, conflict and food insecurity in many settings with fragile food supply chains risk resurgence and perpetuation of childhood malnutrition ^5^.

Understanding long-term outcomes following child malnutrition is especially important because improved treatment has thankfully resulted in more children with malnutrition surviving into adolescence and adulthood ^3, 6^. Most previous research and programmatic investment have focused on child mortality rather than thriving and long-term outcomes. Fewer studies explore long-term impacts of malnutrition on cognitive, behavioural and mental health outcomes in survivors. Improved understanding of these outcomes can inform disease burden estimates, support ongoing investment and inform follow-up care for children with malnutrition.

Causal pathways linking malnutrition with neurodevelopment, cognition, behaviour and mental health are complex. Previous studies have found children admitted to hospital with severe malnutrition often have severe developmental delays and children with neurodisability are inherently at higher risk of becoming malnourished ^7, 8^. The relationship between malnutrition and development is also influenced by other associated factors such as childhood adversity, HIV-exposure, risk of infectious disease, parental engagement, school attendance and impaired growth ^9^. These mediating factors also explain in part the difficulty in predicting developmental trajectories after an episode of severe malnutrition and why studies investigating outcomes are significantly influenced by potential confounders affecting the internal validity of results ^10^.

A 1995 review found school-age children who suffered from early childhood malnutrition generally had poorer IQ levels, cognitive function, school achievement and greater behavioural problems than matched controls and, to a lesser extent, siblings ^11^. Despite these associations, previous evidence of direct causal relationships is limited due to a lack of long-term follow-up studies, retrospective study designs and few studies having investigated behaviour and mental health. With this lack of up-to-date evidence on an increasingly important topic we aimed to identify studies reporting neurodevelopmental, cognitive, behavioural and mental health outcomes for children exposed to malnutrition in childhood.

## METHODS

Our systematic review protocol was registered on PROSPERO (CRD42021260498). We searched MEDLINE, EMBASE, Global Health and PsycINFO for studies published between 1^st^ January 1995 and 6^th^ January 2022 using PRISMA guidelines ^12^. Detailed search strategies are included in Supplementary Appendix 1.

### Selection criteria

We included studies from low- and middle-income countries reporting neurodevelopmental, cognitive, school/academic achievement, mental health or behavioural outcomes in children under five exposed to malnutrition (we did not exclude studies that included small numbers of children between the ages of five and six years). We defined childhood malnutrition as undernutrition using standard definitions (definitions which are commonly measured in research and often related to severe adverse outcomes ^13^): moderate or severe wasting defined by low weight for height or low mid-upper arm circumference; presence of nutritional oedema; low weight-for-age as per older definitions of severe malnutrition. We did not include studies focusing on stunting (low height-for-age), chronic malnutrition or micronutrient deficiencies.

We included cross-sectional, cohort, case control and controlled trial study designs which had a comparator group not exposed to childhood malnutrition. We excluded conference papers and reviews which did not include original data. We excluded studies which failed to define how malnutrition, child neurodevelopment, cognition, school/academic achievement, mental health or behaviour were measured. We also excluded studies looking at specific sub-populations of children (e.g. only children with a specific medical condition). No language restrictions were placed on studies.

### Literature search

Titles and abstracts were screened by two independent reviewers (two of AK, MGo, MKE, MC). Selected full texts by any reviewer were then independently reviewed by two reviewers (two of AK, MGo, MKE, MC) against our selection criteria. We also screened reference lists of included studies. Data extraction of included studies was done by one of AK or MGo into a standard template (including study characteristics, assessment tool, malnutrition definition, sample size, results in cases and controls, reported effect sizes, results of statistical significance tests, results of analyses adjusted for confounding variables). Data extraction was subsequently checked by a second separate reviewer (one of AK, ME, MC). Disagreements regarding inclusion of full texts and data extraction were resolved through mutual discussion.

### Quality assessment

We assessed study quality using the National Institute of Health and Care Excellence (NICE) quality appraisal checklist ^14^. Studies were scored independently by two reviewers (two of AK, MGo, MKE, MC) with scoring disagreements resolved through mutual discussion. Internal and external validity were rated as poor, acceptable and very good based on NICE quality appraisal checklist results. We considered overall study quality to be high-quality when both internal and external validity scores were rated as very good, adequate-quality when either score was rated as acceptable and poor-quality where either internal or external validity was rated as poor.

### Synthesis of study results

We grouped studies into three outcome areas: neurodevelopmental, cognition/academic achievement and mental health/behavioural outcomes (with some studies reporting outcomes from more than one category). We undertook a narrative synthesis of results within each of these areas as diverse outcomes and measurement tools precluded meta-analysis.

We followed the Synthesis without meta-analysis (SWiM) reporting guidelines for analysing and reporting results ^15^. For neurodevelopmental studies we grouped studies and compared results by neurodevelopmental assessment tool used. For studies investigating cognition/academic achievement, we grouped those that measured IQ or executive function and those that measured either school or language performance/assessment results. For mental health/behaviour studies, we grouped studies that used behavioural assessment tools and then into groups by mental health condition or domain assessed.

For each study outcome where available, we recorded the unadjusted and adjusted results in cases and controls, reported effect sizes, and results from any statistical significance tests. When synthesizing results by the groupings described above, we used vote counting by direction of effect to assess the number of studies which recorded differences between cases and controls. We summarised results in tables indicating the number of studies with results indicating an effect of malnutrition on the study outcome within each outcome area. We also recorded the age group (childhood 0-10 years, adolescence 10-18 years, adulthood 18+ years) at which outcomes were measured to compare results between studies. When drawing conclusions from our synthesis we accounted for study quality scoring, whether results adjusted for confounding variables, sample size and the use of validated outcome tools/assessments. Where several high-quality studies in one grouped outcome area reported results of an association with malnutrition, we graded conclusions as strong evidence. Where most adequate and high-quality studies reported an association with malnutrition, but there were uncertainties or limitations of studies identified, we graded conclusions as moderate evidence. Where there were few studies identified or studies with mixed results or poor study quality, we graded associations as inconclusive.

## RESULTS

### Study Characteristics

Thirty studies met our selection criteria (Supplementary Figure 1) ^16–45^. Full study characteristics are included in Supplementary Table 1. Studies were conducted between 1967-2019 across several countries in Africa, Asia and South America and included some long-term cohorts following participants through to adolescence and adulthood. Nine studies were part of the same Barbados Nutrition Study (BNS), a prospective lifelong cohort study assessing multiple outcomes at different follow-up points for children who suffered from malnutrition in the first year of life ^37–45^. Nine studies assessed neurodevelopmental outcomes, 12 studies assessed cognition or academic achievement (three from BNS) and 14 studies assessed mental health conditions or behaviour (eight from BNS). Study designs included cross-sectional, retrospective cohorts, prospective cohorts and randomised controlled trial studies. Neurodevelopmental studies were either cross-sectional or had a short follow-up time during childhood. The age at follow-up following exposure to malnutrition in other studies varied from childhood through to adulthood.

Study quality scoring is included in Supplementary Table 2. Twenty-three studies had very good external validity, with the seven others apart from one with acceptable external validity. This was due to most providing good descriptions of study settings, populations and selection criteria. Fourteen studies had very good internal validity, 12 had acceptable internal validity and four studies had poor internal validity. Studies often scored poorly where important confounding variables such as socioeconomic status weren’t accounted for in analyses, sample size was small or study designs were poor or poorly described, leading to potential biases. Overall, 14 studies were rated as high-quality, 12 were adequate-quality and four were poor-quality.

### Malnutrition and neurodevelopment

We identified nine studies assessing ne3urodevelopmental outcomes of malnutrition (Table 1). A summary of results from these studies is included in Supplementary Table 3. Five studies used the Bayley Scales of Infant Development (BSID). Three of these were adequate-quality studies which found impaired neurodevelopment in those with malnutrition unadjusted for confounding variables such as socioeconomic status and family characteristics ^19, 30, 36^. Two high-quality studies using BSID found impaired neurodevelopment in those with malnutrition but one found that differences were no longer significant when adjusting for differences in current weight during follow-up at two years old ^23, 33^. A further high-quality study using the Denver-II tool found children with malnutrition had lower scores across fine motor, gross motor, language and personal-social domains compared to controls ^22^. One adequate-quality study using a neurodevelopment tool developed in India (Indian Council Medical Research Psychosocial Developmental Screening Test) found poorer development outcomes for those with malnutrition ^34^. Another adequate-quality study using UNICEF Multiple Indicator Cluster Surveys (MICS) Early Childhood Development Indicators (ECDI) did not find an effect of malnutrition on neurodevelopment ^20^. This was carried out on a large population sample from five country surveys in Asia, but the MICS ECDI tool consists of few basic developmental items which span a large age range. This is notably different to the detailed neurodevelopmental assessments by trained assessors used in most other studies. A poor-quality study using an Indian specific development tool (Indian Development Inventory), found poorer neurodevelopment scores in cases, but this was no longer significant when adjusted for confounding variables. The results from high- and adequate-quality studies provides strong evidence of an association between malnutrition in childhood and impaired neurodevelopment across multiple developmental domains.

**Table 1:**
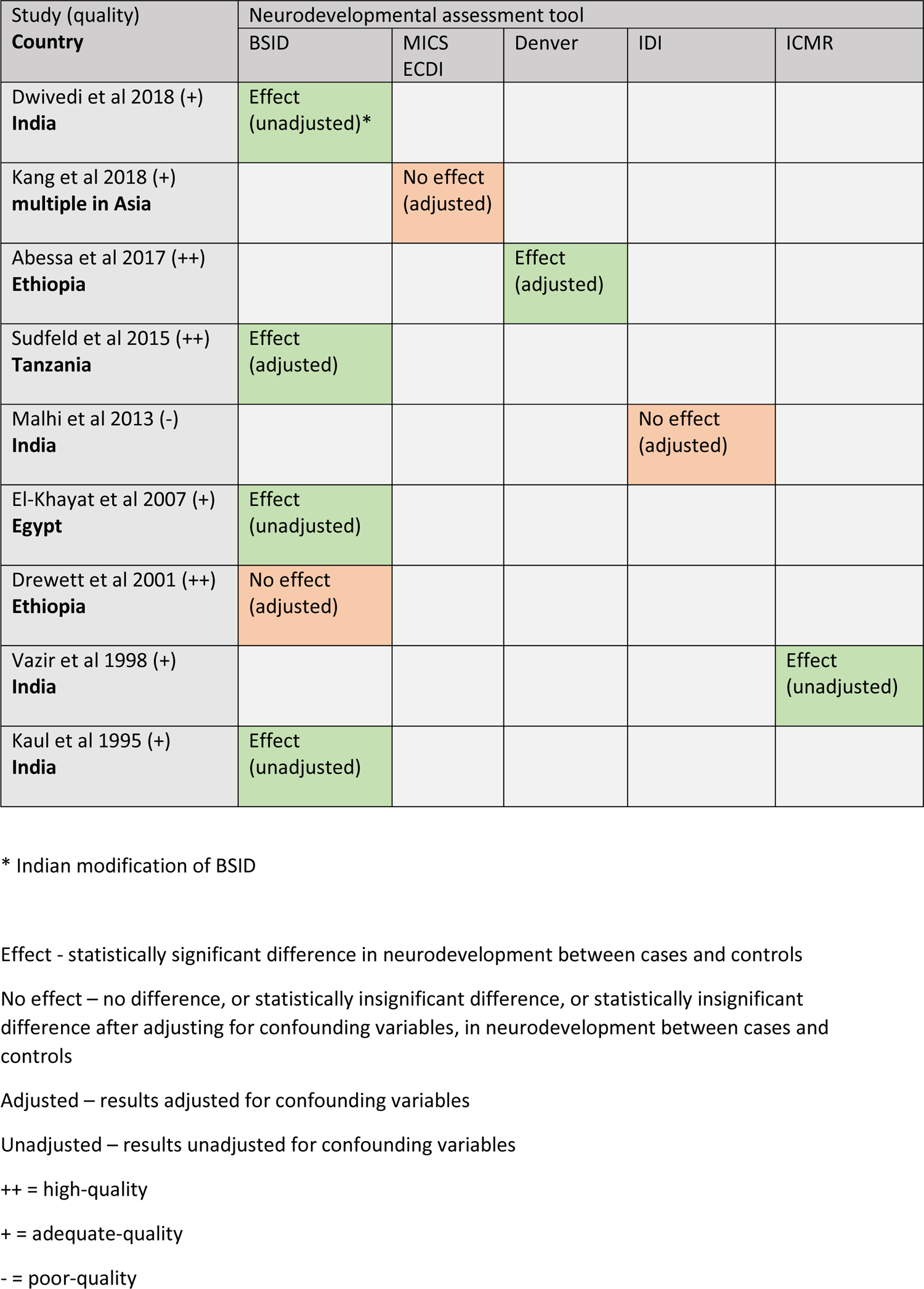
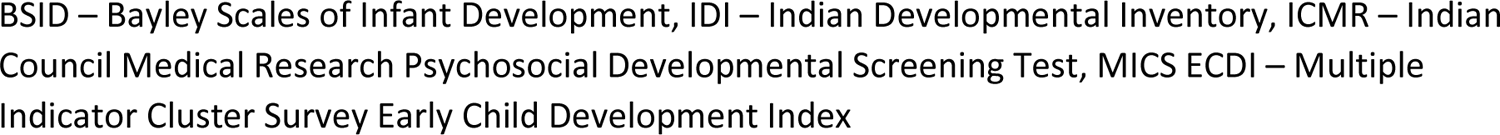
Summary of results from studies assessing the effect of malnutrition on neurodevelopment

### Malnutrition and cognition/academic achievement

We identified 12 studies (three from BNS) assessing cognitive outcomes of malnutrition (Table 2). A summary of results from these studies is included in Supplementary Table 4. Seven studies assessed academic performance, with three based on self-report of school performance or schooling years achieved, and four based on tests of school skill such as mathematics or country specific language tests (Supplementary Table 4) ^17, 18, 21, 28, 32, 38, 43^. Three studies were high-quality, three were adequate-quality and one was poor-quality. All studies (two from BNS) found worse school/academic performance in those exposed to malnutrition in childhood compared to controls. The results from high- and adequate-quality studies identified provides strong evidence of an association between exposure to malnutrition and impaired academic performance/achievement in childhood and adolescence.

**Table 2:**
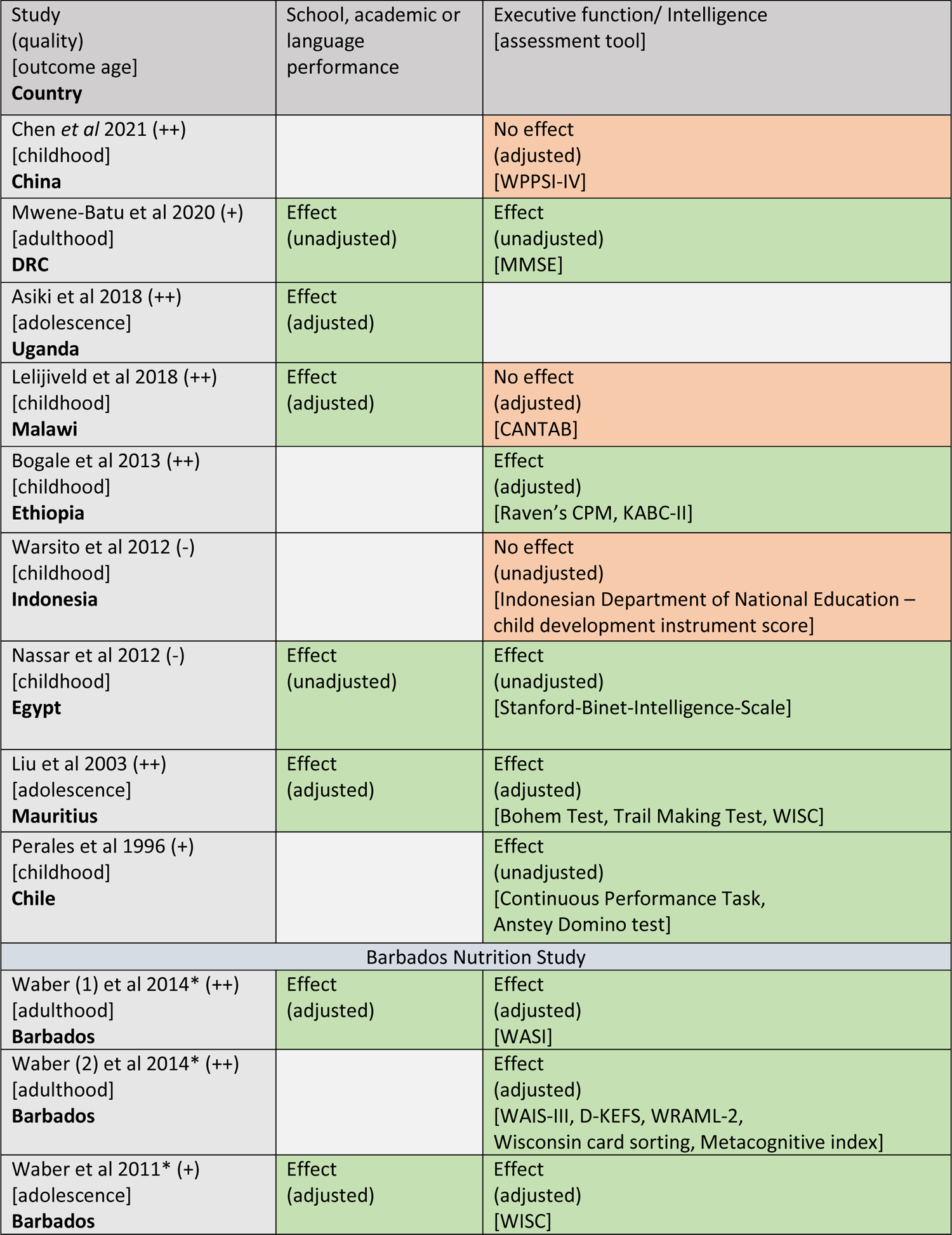
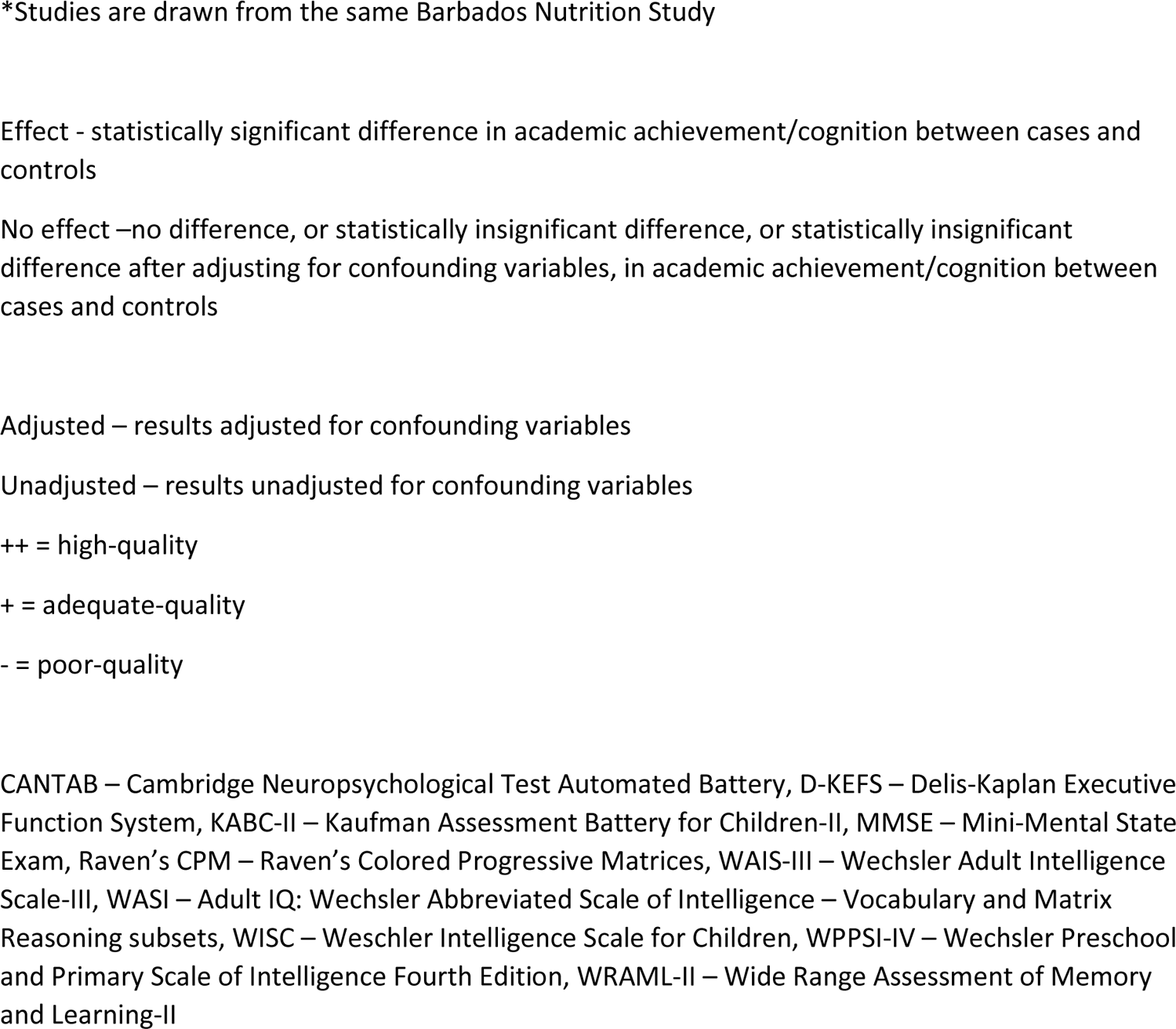
Summary of results from studies assessing the effect of malnutrition on cognition and academic achievement

Eleven studies investigated cognition using several tools assessing intelligence and executive function (Table 2, Supplementary Table 4). Eight of these studies (three from BNS) found impaired intelligence/executive function in those exposed to malnutrition in childhood compared to controls ^17, 26, 28, 32, 35, 38, 39, 43^. Two high-quality studies found no significant association between malnutrition and cognition ^16, 21^. One study from China found no difference between cases and controls using Wechsler Pre-school and Primary Scale of Intelligence (WPPSI-IV), and the other from Malawi found the differences seen between cases and controls using the Cambridge Neuropsychological Test Automated Battery (CANTAB) were no longer significant when adjusting for confounding variables including HIV, stunting, socioeconomic status and household characteristics. Of the studies which found a significant difference in cognition between cases and controls, there were four high quality studies (two from BNS) ^26, 32, 38, 39^. These four high-quality studies all used different cognitive assessment tools, including Kauffman-ABC, Bohem Test, Trail Making Test and Wechsler Scale of Intelligence (Table 2), and found poorer scores in those exposed to malnutrition compared to controls. Three adequate-quality studies (one from BNS) using different cognitive assessment tools, including Mini-Mental State Exam, Wechsler Scale of Intelligence and Anstey Domino test (Table 2) also found those exposed to childhood malnutrition had impaired cognition compared to controls, however only one of these studies adjusted for any confounding variables (BNS study which adjusted for childhood standard of living) ^17, 35, 43^. The results from high- and adequate-quality studies provide moderate evidence of an association between exposure to malnutrition in childhood and impaired cognition, but definitive conclusions are limited by mixed results from two high-quality studies showing no association after adjusting for important confounders such as HIV and socioeconomic status and results from several adequate-quality studies which failed to adjust for any confounding variables.

### Malnutrition and mental health/behaviour

We identified 14 studies (eight from BNS) assessing mental health and behavioural outcomes of malnutrition (Table 3). A summary of results from these studies is included in Supplementary Table 5. Seven studies (three from BNS) assessed behaviour using different behavioural rating tools including the Strengths and Difficulties (SDQ), Child Behaviour Checklist, Ages and Stages (socio-emotional questions) and other adapted behavioural questionnaires (Table 3) ^16, 22, 29, 31, 39, 42, 44^. Five of these studies, four of which were rated as high-quality, found significantly higher behavioural problems in those exposed to malnutrition in childhood ^22, 29, 31, 39, 44^. One high-quality study found no difference in behavioural scores between cases and controls using SDQ and one adequate-quality study looking at conduct problems using found differences in behaviour were no longer significant when adjusting for confounding variables including childhood standard of living. The results from adequate- and high-quality studies provide moderate evidence of an association between exposure to malnutrition in childhood and increased behavioural problems but is limited by some mixed results in high-quality studies and potential confounding in some studies reporting an association.

**Table 3:** Summary of results from studies assessing the effect of malnutrition on mental health and behaviour

| Study<br>(quality)<br>[outcome age]<br>**Country** | Behavioural<br>problems<br>[tool] | Self-esteem<br>[tool] | Social-related<br>disability<br>[tool] | Quality of life<br>[tool] | Personality<br>[tool] | Attention<br>[tool] | Morale<br>[tool] |
| --- | --- | --- | --- | --- | --- | --- | --- |
| Chen *et al* 2021<br>(++)<br>[childhood]<br>**China** | No effect<br>(adjusted)<br>[SDQ] |  |  |  |  |  |  |
| Mwene-Batu et al<br>2020 (+)<br>[adulthood]<br>**DRC** |  | Effect<br>(unadjusted)<br>[Rosenberg Self-Esteem Scale] | No effect<br>(unadjusted)<br>[WHODAS] |  |  |  |  |
| Abessa et al<br>2017 (++)<br>[childhood]<br>**Ethiopia** | Effect<br>(adjusted)<br>[ASQ:SE] |  |  |  |  |  |  |
| De Grandis et al<br>2014 (-)<br>[childhood]<br>**Argentina** |  |  |  | Effect<br>(unadjusted)<br>[Paediatric<br>quality of life<br>inventory] |  |  |  |
| Baker-Henningham et al<br>2009 (++)<br>[childhood]<br>**Bangladesh** | Effect<br>(adjusted)<br>[Wachs<br>temperament<br>questionnaire] |  |  |  |  |  |  |
| Liu et al 2004 (+)<br>[childhood,<br>adolescence]<br>**Mauritius** | Effect<br>(adjusted)<br>[Child Behaviour<br>Checklist] |  |  |  |  |  |  |
| Barbados Nutrition Study |  |  |  |  |  |  |  |
| Hock et al 2018*<br>(+)<br>[adulthood]<br>**Barbados** |  |  |  |  | Mixed<br>(adjusted)<br>[SCID-II-PQ,<br>NEO PI-R FFM] |  |  |
| Waber (2) et al<br>2014* (++)<br>[adulthood]<br>**Barbados** | Effect<br>(adjusted)<br>[Behavioural<br>regulation index] |  |  |  |  |  |  |
| Galler et al<br>2013* (+)<br>[adulthood]<br>**Barbados** |  |  |  |  | Effect<br>(adjusted)<br>[NEO-PI-R] |  |  |
| Galler (1) et al<br>2012* (+)<br>[adulthood]<br>**Barbados** |  |  |  |  |  | Mixed (adjusted)<br>[CAARS, CPT] |  |
| Galler (2) et al<br>2012* (+)<br>[adolescence]<br>**Barbados** | No effect<br>(adjusted)<br>[Conduct: CBQ,<br>Teacher<br>behaviour<br>questionnaire] |  |  |  |  |  |  |
| Waber et al<br>2011* (+)<br>[adolescence]<br>**Barbados** |  |  |  |  |  | No effect<br>(adjusted)<br>[Attention: CBQ] | No effect<br>(adjusted)<br>[Minnesota<br>Adjustment and<br>Morale Scale] |
| Galler et al<br>2011* (++)<br>[adolescence]<br>**Barbados** | Effect<br>(adjusted)<br>[BCBS] |  |  |  |  |  |  |
| Galler et al<br>2010* (++) |  |  |  |  |  |  | Effect |
| [adolescence]<br>**Barbados** |  |  |  |  |  |  | (adjusted)<br>[Minnesota<br>Adjustment and<br>Morale Scale] |

\*Studies are drawn from the same Barbados Nutrition Study
Effect - statistically significant difference in mental health/behavior between cases and controls
No effect –no difference, or statistically insignificant difference, or statistically insignificant difference after adjusting for confounding variables, in mental health/behaviour between cases and controls
Adjusted – results adjusted for confounding variables
Unadjusted – results unadjusted for confounding variables
++ = high-quality
+ = adequate-quality
- = poor-quality
ASQ:SE – Ages and Stages Questionnaire Socio emotional, BCBS – Barbados Child Behaviour Scale, CAARS - Attention-domain: Conners-ADHD-Rating-Scales, CBQ – Child Behaviour Questionnaire, CPT – Conners-Continuous-Performance Test, NEO PI-R – NEO Personality Inventory-Revised, NEO PI-R FFM – NEO Personality Inventory-Revised derived Five-Factor Model, SCID-II-PQ – Structured Clinical Interview for DSM-IV Axis II Personality Disorders Personality Questionnaire, SDQ – Strengths and Difficulties Questionnaire, WHODAS – World Health Organization Disability Assessment Schedule

Seven studies (five from BNS) investigated different mental health outcomes (Table 3) ^17, 24, 37, 40, 41, 43, 45^. Studies indicated possible associations with self-esteem, quality of life, personality disorders, attention deficits and low morale. There were small numbers of studies investigating specific mental health domains and there were significant study limitations in several studies included. The results from the studies identified provide inconclusive evidence regarding possible associations between exposure to malnutrition in childhood and poorer mental health outcomes.

## DISCUSSION

Our review finds strong evidence that exposure to malnutrition in childhood impairs neurodevelopment and academic achievement. There is moderate evidence that childhood malnutrition is associated with impaired cognition and is associated with more behavioural problems throughout childhood and adolescence. However, there is uncertainty around the relative contributions of malnutrition and other associated factors (such as childhood adversity, HIV-exposure, socioeconomic status and household characteristics) to these outcomes. Research investigating mental health outcomes in children with malnutrition is inconclusive and there are few studies investigating specific mental health domains such as depression. These results have implications for policy makers surrounding the long-term care needs of those treated for childhood malnutrition and future research such as exploring how nutritional therapies affect these outcomes.

Due to study heterogeneity, we were unable to perform a meta-analysis of any results. We therefore used published guidelines to synthesize results and determine the strength of evidence for each outcome area. Despite this, there are still limitations given potential publication bias of positive associations between malnutrition and the outcomes investigated. Our review however builds on previously published evidence which has suggested links between malnutrition and poorer IQ levels, cognitive function, school achievement and greater behavioural problems ^11^. Our findings strengthen the evidence-base regarding these associations as previous findings were limited by difficulties in interpreting retrospective case control studies, but our review includes evidence from prospective studies which have sought to minimise these biases. There are however several poor- or adequate-quality studies we identified which failed to account for important confounders such as socioeconomic status and family characteristics. Of the 30 studies we identified, nine were from the same prospective cohort study in Barbados, with many of these publications from the same cohort testing multiple cognitive, behavioural and mental health outcomes. This may have increased the likelihood of type one error. For example, different studies from the same cohort found mixed results regarding morale and personality disorder scores when different assessment tools were used (Table 3) ^41, 43, 45^.

Impaired neurodevelopment in childhood is likely linked to subsequently poorer academic achievement in childhood and adolescence and may also be linked to increased behavioural problems seen in some studies ^46^. The studies assessing IQ/executive function outcomes spanned childhood through to adulthood and there was moderate evidence of a link between malnutrition and cognition. Two high-quality studies showed no effect ^16, 21^ and other studies which showed an effect did not adjust for any confounding variables ^17, 28, 35^. There are therefore outstanding questions over how much early impairment in neurodevelopment due to malnutrition affects future cognition and functioning, and to what extent other related environmental factors such as prenatal nutrition, family characteristics and infections contribute to these outcomes ^47, 48^. Possible sex-specific differences are another area which data here does not allow us to comment on but is important for future work to explore ^49, 50^. Mental health outcomes such as depression and inattention are likely to be influenced by similar confounding variables, and whether early insults from malnutrition independently contribute to poor mental health outcomes in later life is yet to be established. There is also uncertainty around the effect of nutritional therapy on long-term cognition and functioning^51^. However, even if some of the effects we report are due to confounding, evidence is clear from our review that children who experience an episode of malnutrition in childhood are at high risk of poorer development, behaviour and cognition. Policy makers should therefore prioritise targeted support for these vulnerable children and programmes to optimise life chances after recovery.

While there is still uncertainty around the relative contributions of associated medical and social factors, there is increasing evidence from this review that the early impacts of malnutrition are related to worse academic, cognitive and behavioural outcomes compared to well-nourished peers. Preventing and decreasing childhood malnutrition is therefore of key importance to prevent serious long-term neurocognitive issues in affected children, particularly given the ongoing high malnutrition prevalence in many low- and middle-income settings. Further research is needed on how to optimise treatment and to best support ongoing care for survivors to improve outcomes in the long-term.

## Supporting information

PRISMA checklist

## Contributions

**AK, MGo, MGl, MK** conceived the study design. **AK** wrote the main draft of the paper. **AK, MGo, MKE, MC** screened studies against selection criteria, extracted/checked data from studies and quality scored studies. **AK, MGo** analysed and synthesized data. All authors reviewed and contributed to the manuscript.

## Funding

AK is supported by a Wellcome Trust Clinical PhD Programme Fellowship (203919/Z/16/Z).

### Ethics Statement

This study did not receive nor require ethics approval, as it does not involve human & animal participants

## Data Availability

All data produced in the present work are contained in the manuscript

## Acknowledgements

Patients and public were not involved in the design and conduct of this study

## Supplementary Appendix 1 – Search strategies

### EMBASE

1. severe acute malnutrition.ti,ab.
2. acute severe malnutrition.ti,ab.
3. acute malnutrition.ti,ab.
4. (acute adj2 malnutrition).ti,ab.
5. “sever* acute* maln*”.ti,ab.
6. protein-energy malnutrition.ti,ab.
7. protein calorie malnutrition.ti,ab.
8. (protein malnutrition or energy malnutrition or energy-protein malnutrition).ti,ab.
9. “acute* maln*”.ti,ab.
10. “kwas?io?kor*”.ti,ab.
11. “marasm*”.ti,ab.
12. “marasm* kwas?io?kor*”.ti,ab.
13. wasting.ti,ab.
14. “acute wast*”.ti,ab.
15. wasted.ti,ab.
16. (acute adj2 wasting).ti,ab.
17. (severe adj2 wasting).ti,ab.
18. acute undernutrition.ti,ab.
19. (acute adj2 undernutrition).ti,ab.
20. severe undernutrition.ti,ab.
21. (severe adj2 undernutrition).ti,ab.
22. “mid* upper arm circumference”.ti,ab.
23. (mid* upper arm circumference adj “115”).ti,ab.
24. (mid* upper arm circumference adj “110”).ti,ab.
25. MUAC.ti,ab.
26. “acute* undernourish*”.ti,ab.
27. “acut* emaciat*”.ti,ab.
28. “acut* maln*”.ti,ab.
29. *malnutrition/
30. exp kwashiorkor/
31. exp marasmus/
32. exp wasting syndrome/
33. exp protein calorie malnutrition/
34. *nutritional disorder/
35. 1 or 2 or 3 or 4 or 5 or 6 or 7 or 8 or 9 or 10 or 11 or 12 or 13 or 14 or 15 or 16 or 17 or 18 or 19 or 20 or 21 or 22 or 23 or 24 or 25 or 26 or 27 or 28 or 29 or 30 or 31 or 32 or 33 or 34
36. infant/
37. child/
38. exp toddler/
39. “child*”.ti,ab.
40. “infan*”.ti,ab.
41. “pre-school*”.ti,ab.
42. “preschool*”.ti,ab.
43. “toddler*”.ti,ab.
44. 36 or 37 or 38 or 39 or 40 or 41 or 42 or 43
45. 35 and 44
46. “child* developm* “.ti,ab.
47. “child* neurodevelopm*”.ti,ab.
48. “neurodevelopm*”.ti,ab.
49. (socio-emotion* develop* or socioemotion* develop*).ti,ab.
50. “social developm*”.ti,ab.
51. “emotion* developm*”.ti,ab.
52. (sensorymotor developm* or sensory-motor developm*).ti,ab.
53. “motor developm*”.ti,ab.
54. “motor neurodevelopm*”.ti,ab.
55. exp child development/ or exp postnatal development/ 56. 46 or 47 or 48 or 49 or 50 or 51 or 52 or 53 or 54 or 55
56. exp cognition assessment/ or exp cognition/
57. exp learning/ or exp learning disorder/
58. exp intelligence quotient/ or exp intelligence/
59. exp language development/ or exp human development/ or exp mental development/
60. *brain function/ or *central nervous system function/
61. cognition.ti,ab.
62. “cogniti* performance”.ti,ab.
63. intelligence.ti,ab.
64. IQ.ti,ab.
65. “executive function*”.ti,ab.
66. reasoning.ti,ab.
67. language.ti,ab.
68. attention.ti,ab.
69. memory.ti,ab.
70. memory.ti,ab.
71. learning.ti,ab.
72. early learning.ti,ab.
73. information processing.ti,ab.
74. literacy.ti,ab.
75. reading.ti,ab.
76. math.ti,ab.
77. 57 or 58 or 59 or 60 or 61 or 62 or 63 or 64 or 65 or 66 or 67 or 69 or 70 or 71 or 72 or 73 or 74 or 75 or 76 or 77
78. education.ti,ab.
79. “school* achievement”.ti,ab.
80. “school* performance”.ti,ab.
81. school retention.ti,ab.
82. academic.ti,ab.
83. pre-academic.ti,ab.
84. schooling.ti,ab.
85. exp academic achievement/
86. 79 or 80 or 81 or 82 or 83 or 84 or 85 or 86
87. exp behavior/
88. exp mental health/
89. exp psychological aspect/ or exp neuropsychology/ or exp attention/ or exp cognitive defect/ or exp mental disease/
90. exp depression/
91. *hyperactivity/ or *psychomotor disorder/
92. exp attention deficit disorder/
93. “behavi?r* problem*”.ti,ab.
94. “emotion* problem*”.ti,ab.
95. “temperament*”.ti,ab.
96. self regulation.ti,ab.
97. attachment.ti,ab.
98. self esteem.ti,ab.
99. self efficacy.ti,ab.
100. “social competen*”.ti,ab.
101. “peer relationship*”.ti,ab.
102. pro-social behavi?r.ti,ab.
103. hyperactivity.ti,ab.
104. impulsivity.ti,ab.
105. attention* deficit hyperactivity disorder*.ti,ab.
106. agression.ti,ab.
107. 88 or 89 or 90 or 91 or 92 or 93 or 94 or 95 or 96 or 97 or 98 or 99 or 100 or 101 or 102 or 103 or 104 or 105 or 106 or 107
108. 56 or 78 or 90 or 108
109. 45 and 109
110. limit 110 to (human and yr=”1995-Current”)

### MEDLINE

1. “acute severe malnutrition”.ti,ab.
2. severe acute malnutrition.ti,ab.
3. acute malnutrition.ti,ab.
4. (acute adj2 malnutrition).ti,ab.
5. “sever* acute* maln*”.ti,ab.
6. protein-energy malnutrition.ti,ab.
7. acute severe malnutrition.ti,ab.
8. (protein malnutrition or energy malnutrition or energy-protein malnutrition).ti,ab.
9. “acute* maln*”.ti,ab.
10. “kwas?io?kor*”.ti,ab.
11. “marasm*”.ti,ab.
12. “marasm* kwas?io?kor*”.ti,ab.
13. wasting.ti,ab.
14. “acute wast*”.ti,ab.
15. wasted.ti,ab.
16. (acute adj2 wasting).ti,ab.
17. (severe adj2 wasting).ti,ab.
18. acute undernutrition.ti,ab.
19. (acute adj2 undernutrition).ti,ab.
20. severe undernutrition.ti,ab.
21. (severe adj2 undernutrition).ti,ab.
22. “mid* upper arm circumference”.ti,ab.
23. (mid* upper arm circumference adj “115”).ti,ab.
24. (mid* upper arm circumference adj “110”).ti,ab.
25. MUAC.ti,ab.
26. “acute* undernourish*”.ti,ab.
27. “acut* maln*”.ti,ab.
28. exp child nutrition disorders/ or exp deficiency diseases/ or exp starvation/ or *wasting syndrome/
29. exp Protein-Energy Malnutrition/
30. exp Infant Nutrition Disorders/
31. 1 or 2 or 3 or 4 or 5 or 6 or 7 or 8 or 9 or 10 or 11 or 12 or 13 or 14 or 15 or 16 or 17 or 18 or 19 or 20 or 21 or 22 or 23 or 24 or 25 or 26 or 27 or 28 or 29 or 30
32. “child*”.ti,ab.
33. “infan*”.ti,ab.
34. “pre-school*”.ti,ab.
35. “preschool*”.ti,ab.
36. “toddler*”.ti,ab.
37. exp Infant/
38. exp Child/
39. Child, Preschool/
40. 32 or 33 or 34 or 35 or 36 or 37 or 38 or 39
41. 31 and 40
42. “child* developm* “.ti,ab.
43. “child* neurodevelopm*”.ti,ab.
44. “neurodevelopm*”.ti,ab.
45. (socio-emotion* develop* or socioemotion* develop*).ti,ab.
46. “social developm*”.ti,ab.
47. “emotion* developm*”.ti,ab.
48. (sensorymotor developm* or sensory-motor developm*).ti,ab.
49. “motor developm*”.ti,ab.
50. “motor neurodevelopm*”.ti,ab.
51. exp Child Development/
52. 42 or 43 or 44 or 45 or 46 or 47 or 48 or 49 or 50 or 51
53. cognition.ti,ab.
54. “cogniti* performance”.ti,ab.
55. intelligence.ti,ab.
56. IQ.ti,ab.
57. “executive function*”.ti,ab.
58. reasoning.ti,ab.
59. language.ti,ab.
60. attention.ti,ab.
61. memory.ti,ab.
62. learning.ti,ab.
63. early learning.ti,ab.
64. information processing.ti,ab.
65. literacy.ti,ab.
66. reading.ti,ab.
67. math.ti,ab.
68. *cognition/ or exp cognitive reserve/ or exp comprehension/ or exp executive function/ or exp learning/
69. exp Mild Cognitive Impairment/
70. *Intelligence/
71. *language development/ or *child language/
72. 53 or 54 or 55 or 56 or 57 or 58 or 59 or 60 or 61 or 62 or 63 or 64 or 65 or 66 or 67 or 68 or 69 or 70 or 71
73. education.ti,ab.
74. “school* achievement”.ti,ab.
75. “school* performance”.ti,ab.
76. school retention.ti,ab.
77. academic.ti,ab.
78. pre-academic.ti,ab.
79. schooling.ti,ab.
80. exp Educational Status/
81. 73 or 74 or 75 or 76 or 77 or 78 or 79 or 80
82. “behavi?r* problem*”.ti,ab.
83. “emotion* problem*”.ti,ab.
84. “temperament*”.ti,ab.
85. self regulation.ti,ab.
86. attachment.ti,ab.
87. self esteem.ti,ab.
88. self efficacy.ti,ab.
89. “social competen*”.ti,ab.
90. “peer relationship*”.ti,ab.
91. pro-social behavi?r.ti,ab.
92. hyperactivity.ti,ab.
93. impulsivity.ti,ab.
94. attention* deficit hyperactivity disorder*.ti,ab.
95. aggression.ti,ab.
96. exp Child Behavior Disorders/
97. exp Mental Health/
98. *psychology/ or exp psychology, adolescent/ or exp psychology, child/
99. exp Depression/
100. exp Attention Deficit Disorder with Hyperactivity/
101. *Anxiety Disorders/
102. 82 or 83 or 84 or 85 or 86 or 87 or 88 or 89 or 90 or 91 or 92 or 93 or 94 or 95 or 96 or 97 or 98 or 99 or 100 or 101 103. 52 or 72 or 81 or 102
103. 41 and 103
104. limit 104 to (humans and yr=”1995-Current”)

### GLOBAL HEALTH

1. severe acute malnutrition.ti,ab.
2. acute severe malnutrition.ti,ab.
3. acute malnutrition.ti,ab.
4. (acute adj2 malnutrition).ti,ab.
5. “sever* acute* maln*”.ti,ab.
6. protein-energy malnutrition.ti,ab.
7. protein calorie malnutrition.ti,ab.
8. (protein malnutrition or energy malnutrition or energy-protein malnutrition).ti,ab.
9. “acute* maln*”.ti,ab.
10. “kwas?io?kor*”.ti,ab.
11. “marasm*”.ti,ab.
12. “marasm* kwas?io?kor*”.ti,ab.
13. wasting.ti,ab.
14. “acute wast*”.ti,ab.
15. wasted.ti,ab.
16. (acute adj2 wasting).ti,ab.
17. (severe adj2 wasting).ti,ab.
18. acute undernutrition.ti,ab.
19. (acute adj2 undernutrition).ti,ab.
20. severe undernutrition.ti,ab.
21. (severe adj2 undernutrition).ti,ab.
22. “mid* upper arm circumference”.ti,ab.
23. (mid* upper arm circumference adj “115”).ti,ab.
24. (mid* upper arm circumference adj “110”).ti,ab.
25. MUAC.ti,ab.
26. “acute* undernourish*”.ti,ab.
27. “acut* maln*”.ti,ab.
28. emaciat*.ti,ab.
29. nutritional disorders/ or malnutrition/ or exp protein energy malnutrition/ or exp undernutrition/ or exp nutritional oedema/
30. wasting disease/ or exp emaciation/
31. 1 or 2 or 3 or 4 or 5 or 6 or 7 or 8 or 9 or 10 or 11 or 12 or 13 or 14 or 15 or 16 or 17 or 18 or 19 or 20 or 21 or 22 or 23 or 24 or 25 or 26 or 27 or 28 or 29 or 30
32. “child*”.ti,ab.
33. “infan*”.ti,ab.
34. “pre-school*”.ti,ab.
35. “preschool*”.ti,ab.
36. “toddler*”.ti,ab.
37. exp infants/
38. exp preschool children/ or children/
39. 32 or 33 or 34 or 35 or 36 or 37 or 38
40. 31 and 39
41. “child* developm* “.ti,ab.
42. “child* neurodevelopm*”.ti,ab.
43. “neurodevelopm*”.ti,ab.
44. (socio-emotion* develop* or socioemotion* develop*).ti,ab.
45. “social developm*”.ti,ab.
46. “emotion* developm*”.ti,ab.
47. (sensorymotor developm* or sensory-motor developm*).ti,ab.
48. “motor developm*”.ti,ab.
49. child development/ or exp early childhood development/ or exp infant development/ or exp psychomotor development/
50. 41 or 42 or 43 or 44 or 45 or 46 or 47 or 48 or 49
51. cognition.ti,ab.
52. “cogniti* performance”.ti,ab.
53. intelligence.ti,ab.
54. IQ.ti,ab.
55. “executive function*”.ti,ab.
56. reasoning.ti,ab.
57. language.ti,ab.
58. attention.ti,ab.
59. memory.ti,ab.
60. learning.ti,ab.
61. early learning.ti,ab.
62. information processing.ti,ab.
63. literacy.ti,ab.
64. reading.ti,ab.
65. math.ti,ab.
66. exp mental ability/ or exp cognitive development/ or exp memory/
67. exp learning/
68. 51 or 52 or 53 or 54 or 55 or 56 or 57 or 58 or 59 or 60 or 61 or 62 or 63 or 64 or 65 or 66 or 67
69. education.ti,ab.
70. “school* achievement”.ti,ab.
71. “school* performance”.ti,ab.
72. school retention.ti,ab.
73. academic.ti,ab.
74. schooling.ti,ab.
75. preacademic.ti,ab.
76. education/ or exp early childhood education/ or exp elementary education/ or exp extension education/ or exp higher education/ or exp primary education/ or exp professional education/ or exp secondary education/ or exp academic achievement/ or exp educational performance/
77. 69 or 70 or 71 or 72 or 73 or 74 or 75 or 76
78. “behavi?r* problem*”.ti,ab.
79. “emotion* problem*”.ti,ab.
80. “temperament*”.ti,ab.
81. self regulation.ti,ab.
82. attachment.ti,ab.
83. self esteem.ti,ab.
84. self efficacy.ti,ab.
85. “social competen*”.ti,ab.
86. “peer relationship*”.ti,ab.
87. pro-social behavi?r.ti,ab.
88. hyperactivity.ti,ab.
89. impulsivity.ti,ab.
90. attention* deficit hyperactivity disorder*.ti,ab.
91. aggression.ti,ab.
92. exp mental health/ or exp mental disorders/
93. depression/
94. psychology/ or exp adolescent development/ or exp adult development/ or exp emotional development/ or exp self esteem/ or exp self reliance/
95. exp attention deficit hyperactivity disorder/
96. 78 or 79 or 80 or 81 or 82 or 83 or 84 or 85 or 86 or 87 or 88 or 89 or 90 or 91 or 92 or 93 or 94 or 95 97. 50 or 68 or 77 or 96
97. 40 and 97
98. limit 98 to yr=”1995-Current”

### PsycINFO

1. severe acute malnutrition.ti,ab.
2. acute malnutrition.ti,ab.
3. (acute adj2 malnutrition).ti,ab.
4. “sever* acute* maln*”.ti,ab.
5. protein-energy malnutrition.ti,ab.
6. protein calorie malnutrition.ti,ab.
7. (protein malnutrition or energy malnutrition or energy-protein malnutrition).ti,ab.
8. “acute* maln*”.ti,ab.
9. “kwas?io?kor*”.ti,ab.
10. “marasm*”.ti,ab.
11. “marasm* kwas?io?kor*”.ti,ab.
12. wasting.ti,ab.
13. wasted.ti,ab.
14. (acute adj2 wasting).ti,ab.
15. (severe adj2 wasting).ti,ab.
16. acute undernutrition.ti,ab.
17. (acute adj2 undernutrition).ti,ab.
18. severe undernutrition.ti,ab.
19. (severe adj2 undernutrition).ti,ab.
20. “mid* upper arm circumference”.ti,ab.
21. MUAC.ti,ab.
22. “acute* undernourish*”.ti,ab.
23. “emaciat*”.ti,ab.
24. “acut* maln*”.ti,ab.
25. nutritional deficiencies/ or protein deficiency disorders/ or exp failure to thrive/ or exp underweight/
26. exp Kwashiorkor/
27. *protein deficiency disorders/
28. 1 or 2 or 3 or 4 or 5 or 6 or 7 or 8 or 9 or 10 or 11 or 12 or 13 or 14 or 15 or 16 or 17 or 18 or 19 or 20 or 21 or 22 or 23 or 24 or 25 or 26 or 27
29. “child*”.ti,ab.
30. “infan*”.ti,ab.
31. “pre-school*”.ti,ab.
32. “preschool*”.ti,ab.
33. “toddler*”.ti,ab.
34. 29 or 30 or 31 or 32 or 33
35. 28 and 34
36. “child* developm* “.ti,ab.
37. “child* neurodevelopm*”.ti,ab.
38. “neurodevelopm*”.ti,ab.
39. (socio-emotion* develop* or socioemotion* develop*).ti,ab.
40. “social developm*”.ti,ab.
41. “emotion* developm*”.ti,ab.
42. (sensorymotor developm* or sensory-motor developm*).ti,ab.
43. “motor developm*”.ti,ab.
44. “motor neurodevelopm*”.ti,ab.
45. psychomotor developm*.ti,ab.
46. exp infant development/ or exp early childhood development/
47. *childhood development/ or exp early childhood development/ or exp motor development/ or exp psychological development/ or exp psychomotor development/
48. exp emotional development/ or exp psychological development/
49. 36 or 37 or 38 or 39 or 40 or 41 or 42 or 43 or 44 or 45 or 46 or 47 or 48
50. cognition.ti,ab.
51. “cogniti* performance”.ti,ab.
52. intelligence.ti,ab.
53. IQ.ti,ab.
54. “executive function*”.ti,ab.
55. reasoning.ti,ab.
56. language.ti,ab.
57. attention.ti,ab.
58. memory.ti,ab.
59. learning.ti,ab.
60. early learning.ti,ab.
61. information processing.ti,ab.
62. literacy.ti,ab.
63. reading.ti,ab.
64. math.ti,ab.
65. *cognitive processes/ or exp cognitive assessment/ or exp learning/ or exp memory/ or exp neurocognition/
66. *learning/ or adult learning/ or problem based learning/ or school learning/ or skill learning/ or verbal learning/
67. exp Fine Motor Skill Learning/ or exp Learning Disabilities/ or exp Learning Disorders/ or exp Gross Motor Skill Learning/
68. *language disorders/ or *specific language impairment/ or exp language delay/ or exp language development/
69. *cognitive development/ or exp brain development/ or exp speech development/
70. *cognition/ or exp cognitive impairment/
71. *intelligence/ or exp intellectual development/ or exp intelligence quotient/
72. 50 or 51 or 52 or 53 or 54 or 55 or 56 or 57 or 58 or 59 or 60 or 61 or 62 or 63 or 64 or 65 or 66 or 67 or 68 or 69 or 70 or 71
73. education.ti,ab.
74. “school* achievement”.ti,ab.
75. “school* performance”.ti,ab.
76. school retention.ti,ab.
77. academic.ti,ab.
78. pre-academic.ti,ab.
79. schooling.ti,ab.
80. *education/
81. education/ or exp academic achievement/ or exp school attendance/ or exp school dropouts/ or exp school learning/
82. 73 or 74 or 75 or 76 or 77 or 78 or 79 or 80 or 81
83. “behavi?r* problem*”.ti,ab.
84. “emotion* problem*”.ti,ab.
85. “temperament*”.ti,ab.
86. self regulation.ti,ab.
87. attachment.ti,ab.
88. self esteem.ti,ab.
89. self efficacy.ti,ab.
90. “social competen*”.ti,ab.
91. “peer relationship*”.ti,ab.
92. pro-social behavi?r.ti,ab.
93. hyperactivity.ti,ab.
94. impulsivity.ti,ab.
95. attention* deficit hyperactivity disorder*.ti,ab.
96. aggression.ti,ab.
97. *mental health/
98. exp behavior problems/ or exp behavior disorders/ or exp conduct disorder/
99. exp anxiety disorders/
100. exp hyperkinesis/ or exp attention deficit disorder with hyperactivity/
101. *deprivation/ or exp psychological stress/ or exp stress/
102. *affective disorders/ or *major depression/
103. 83 or 84 or 85 or 86 or 87 or 88 or 89 or 90 or 91 or 92 or 93 or 94 or 95 or 96 or 97 or 98 or 99 or 100 or 101 or 102
104. 49 or 72 or 82 or 103
105. 35 and 104
106. limit 105 to (human and yr=”1995 –Current)

---

**Supplementary Figure 1.**
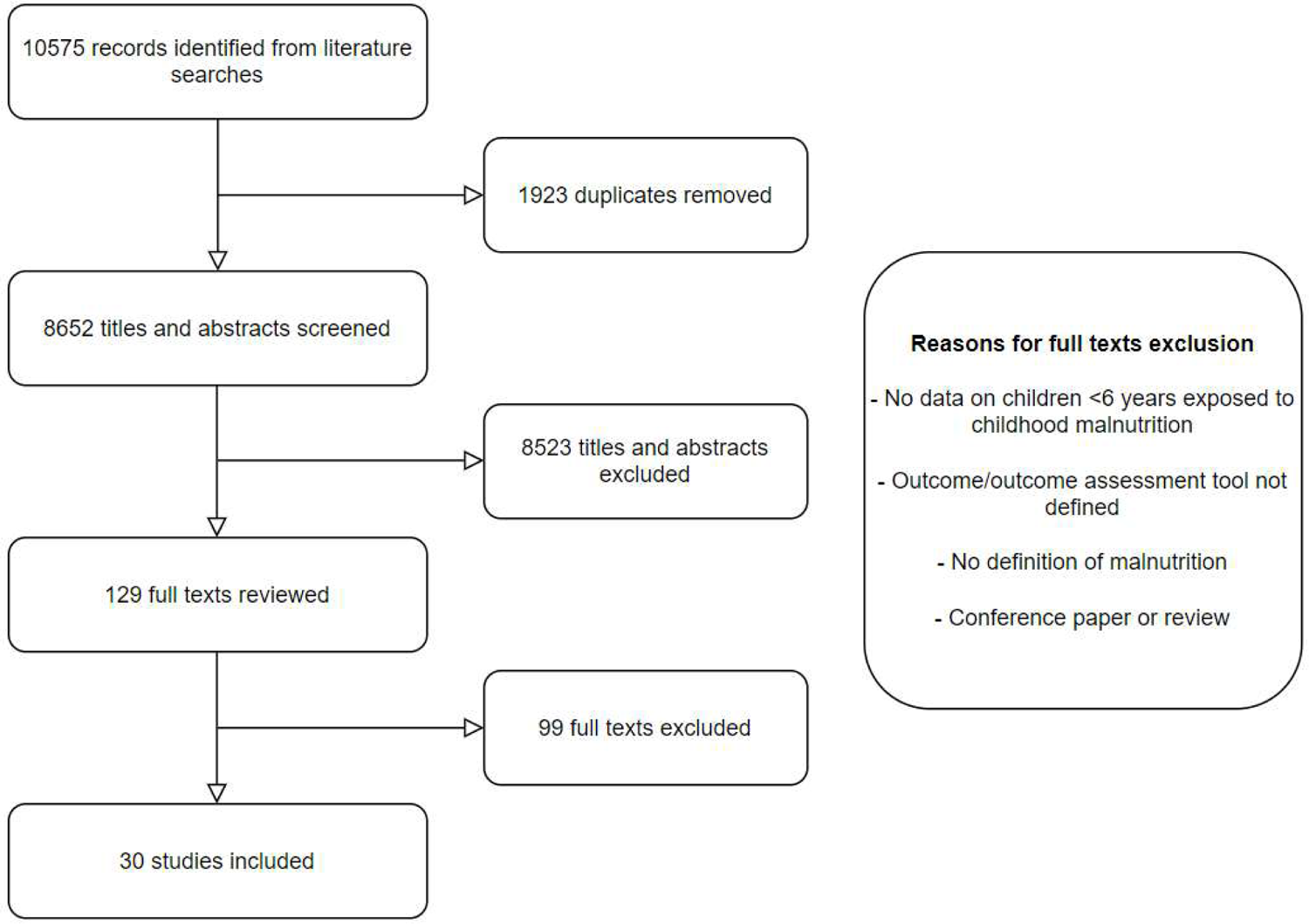
Flow chart of literature search

**Supplementary Table 1.**
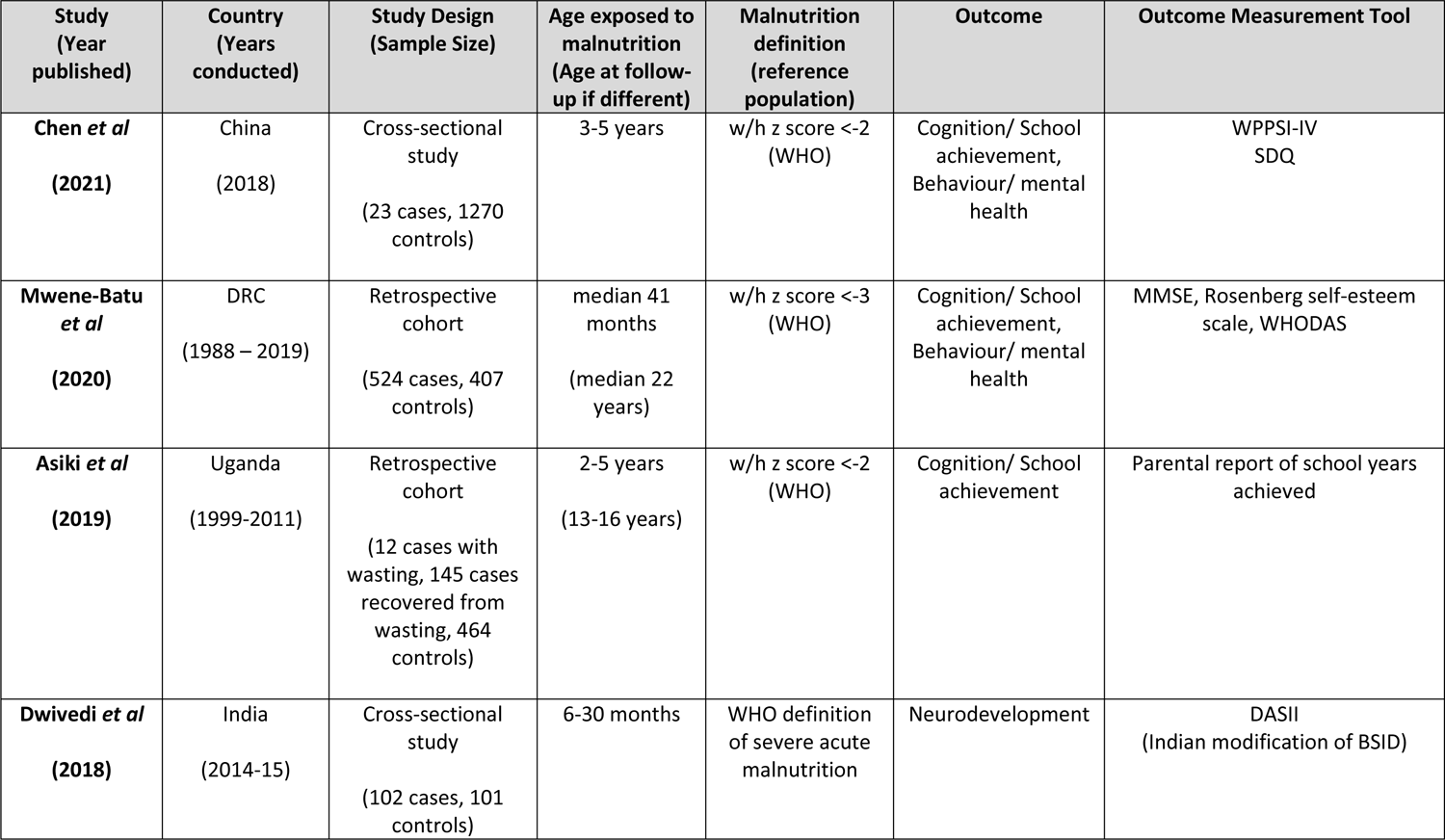
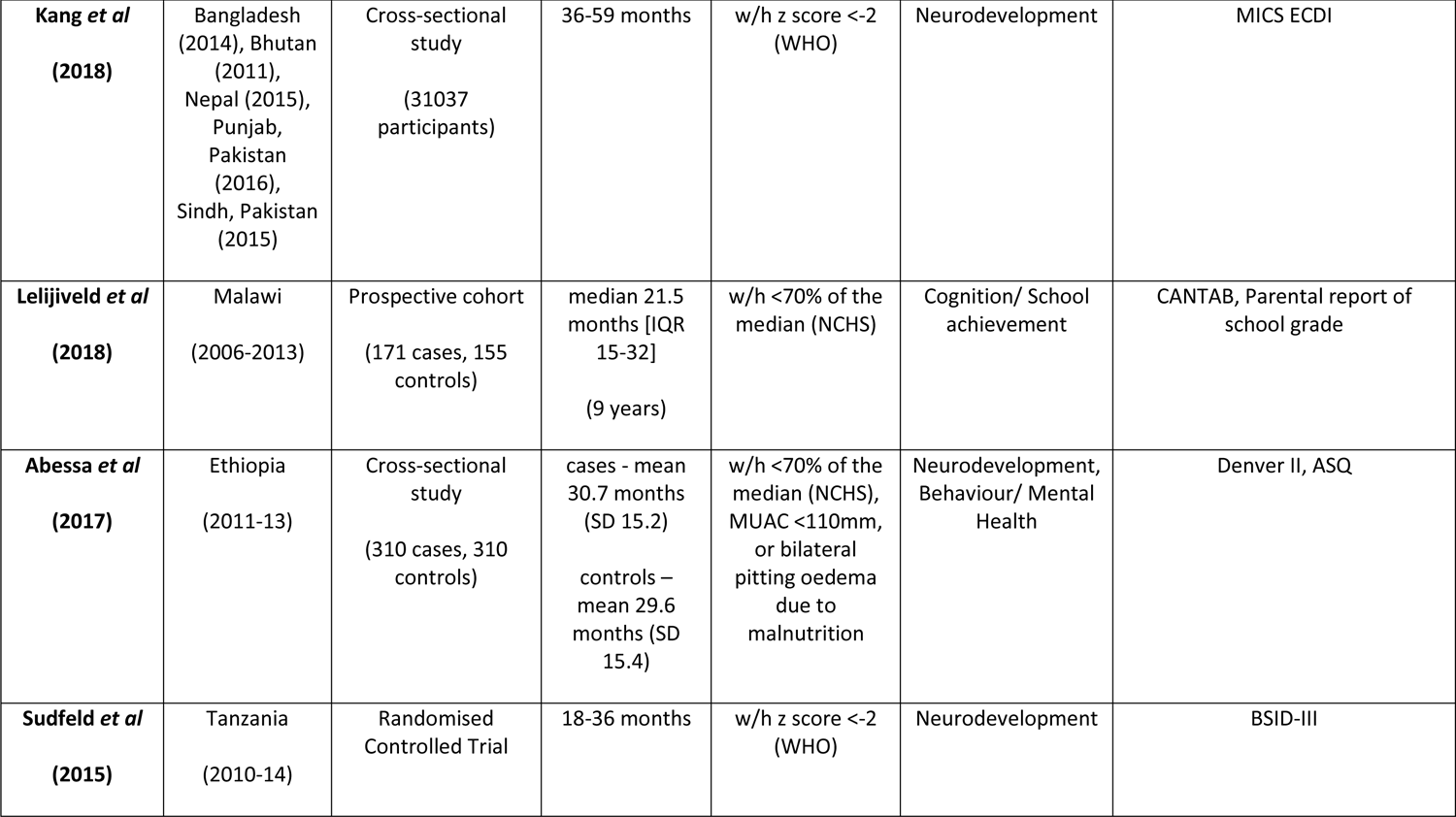
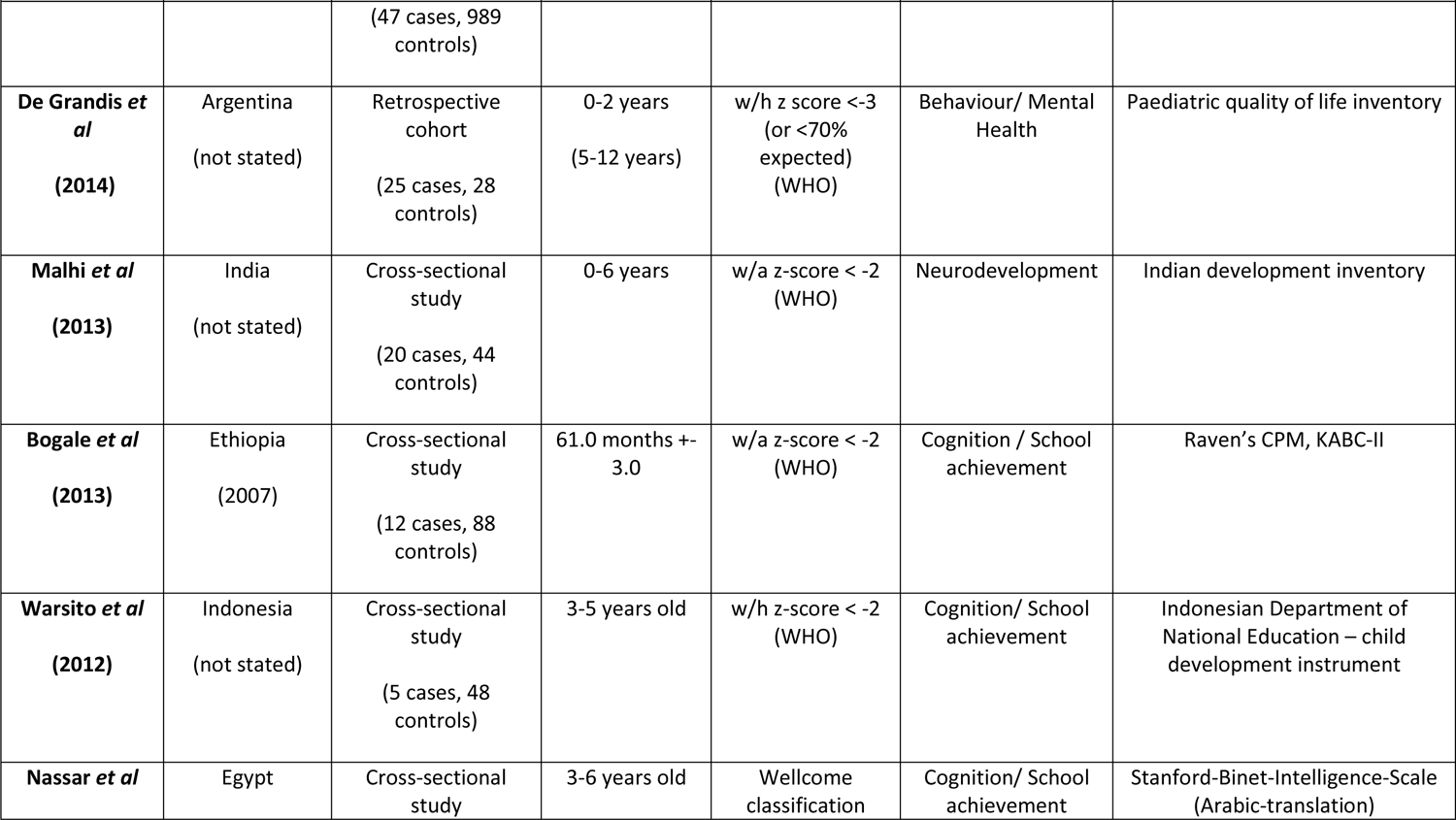
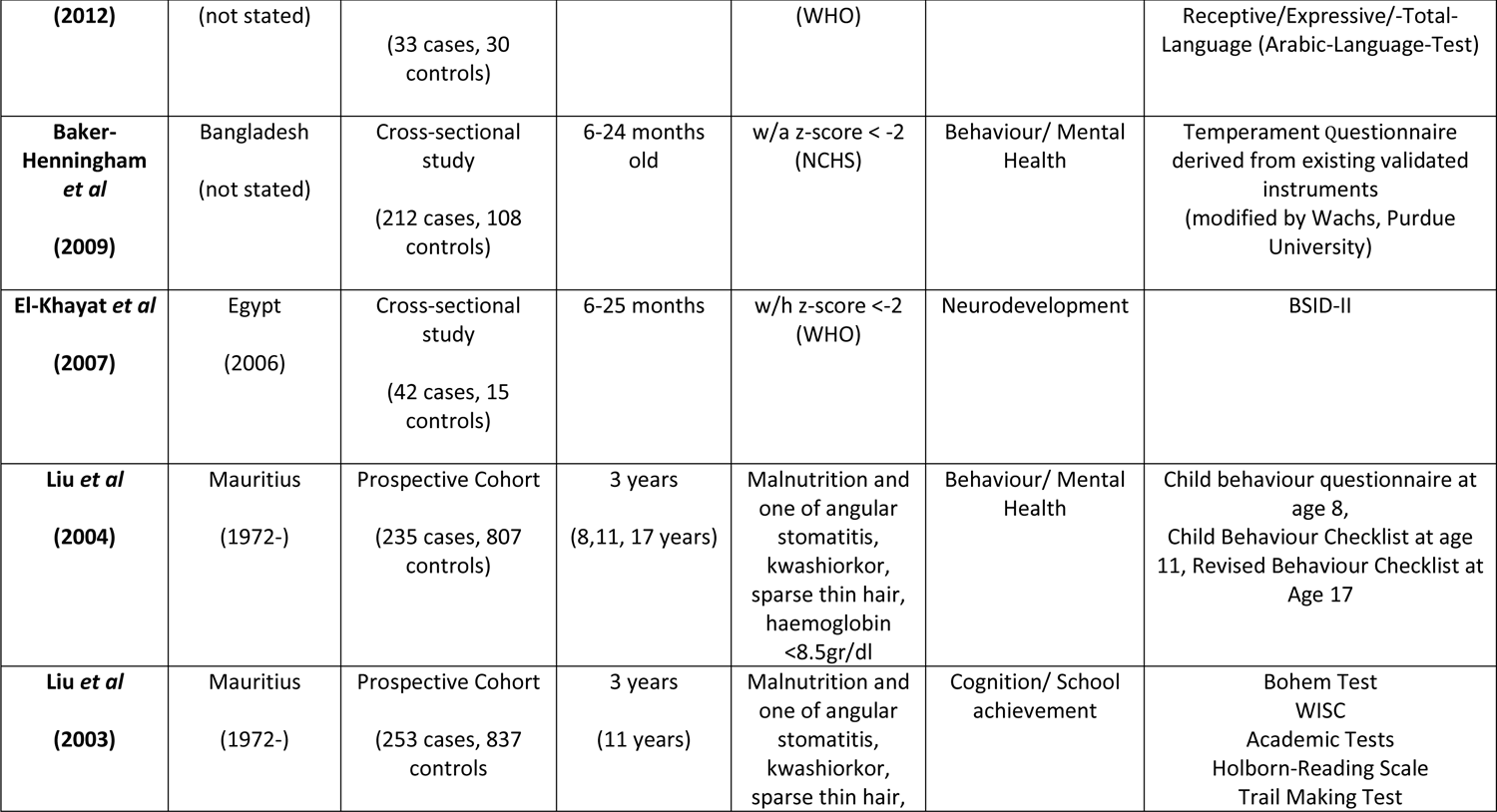
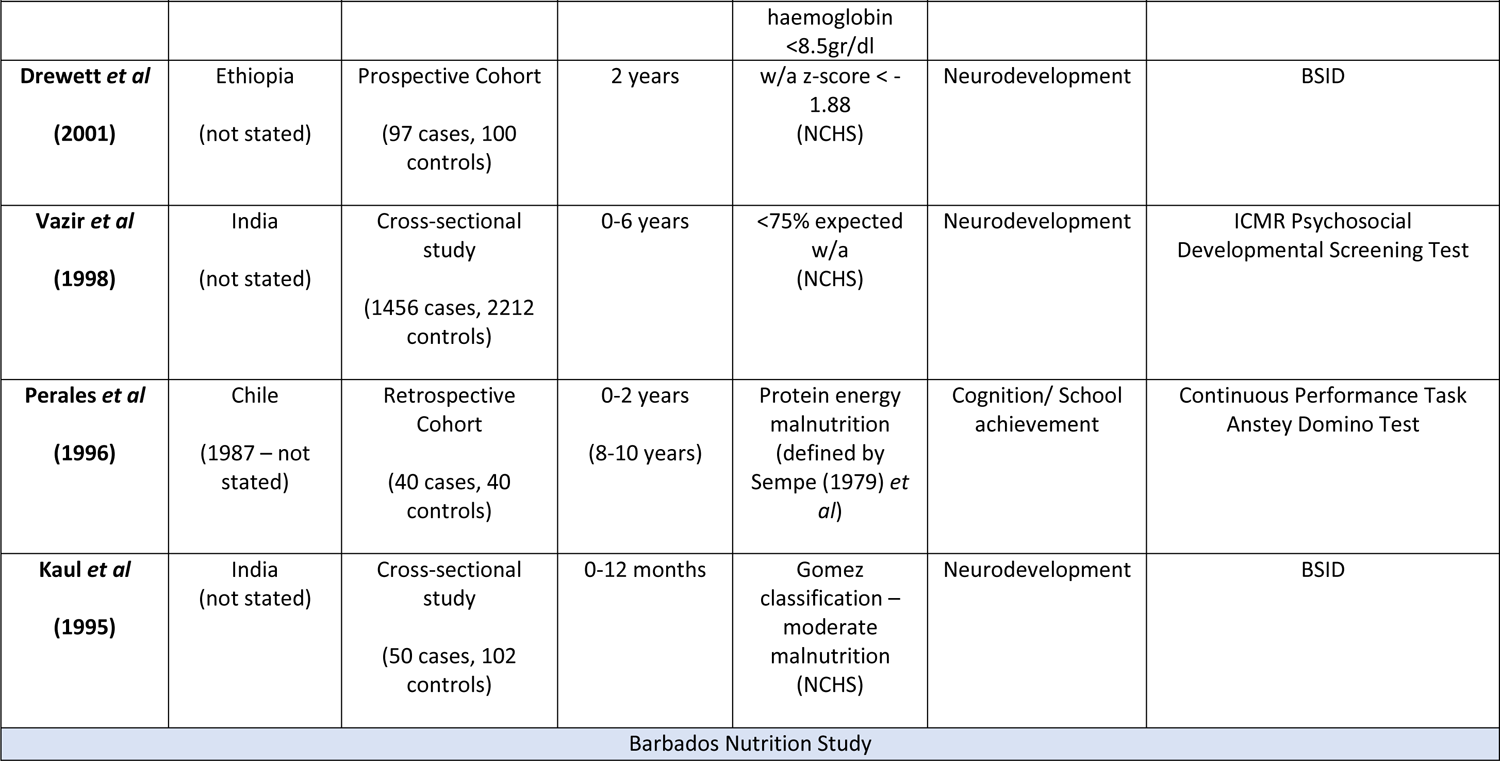
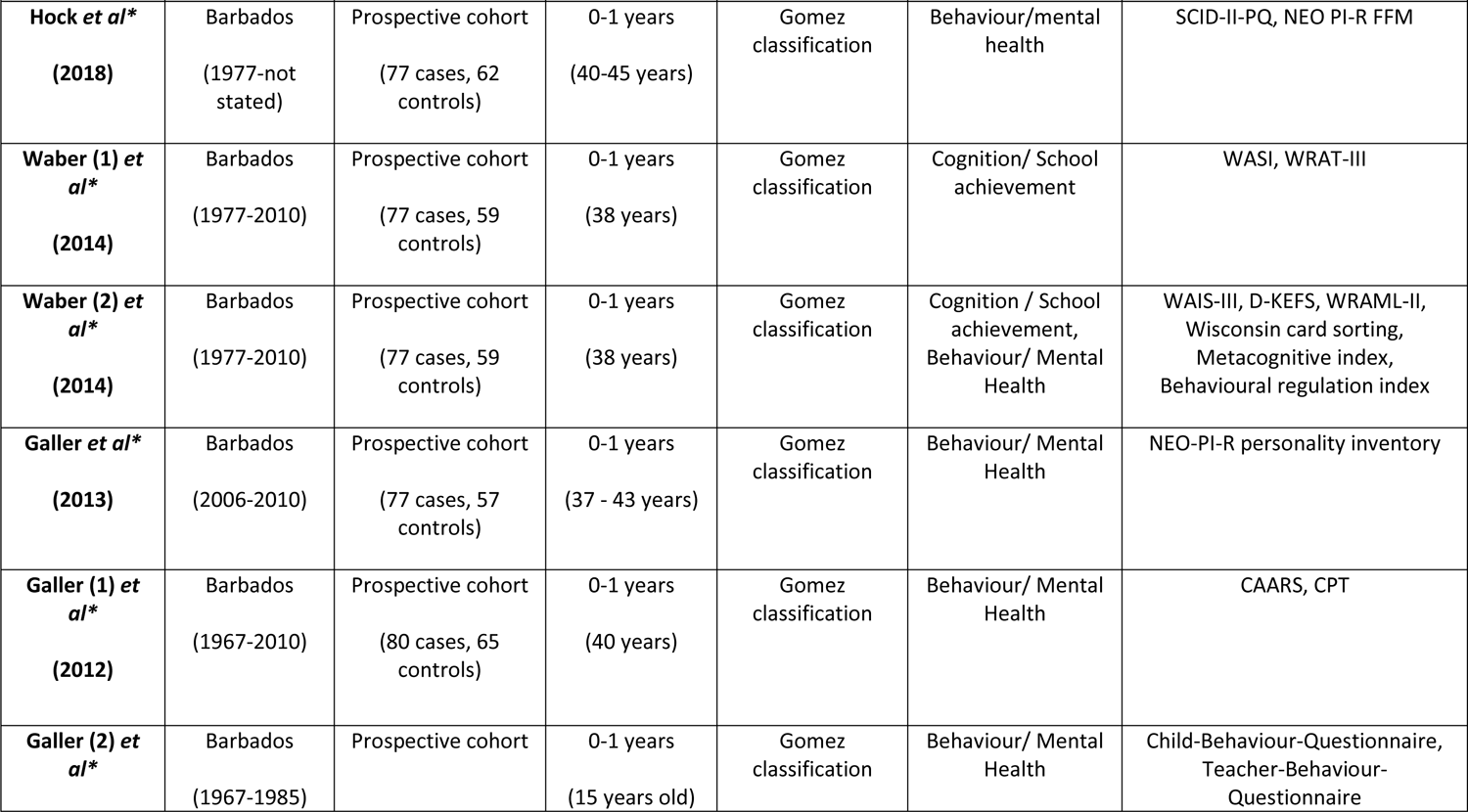
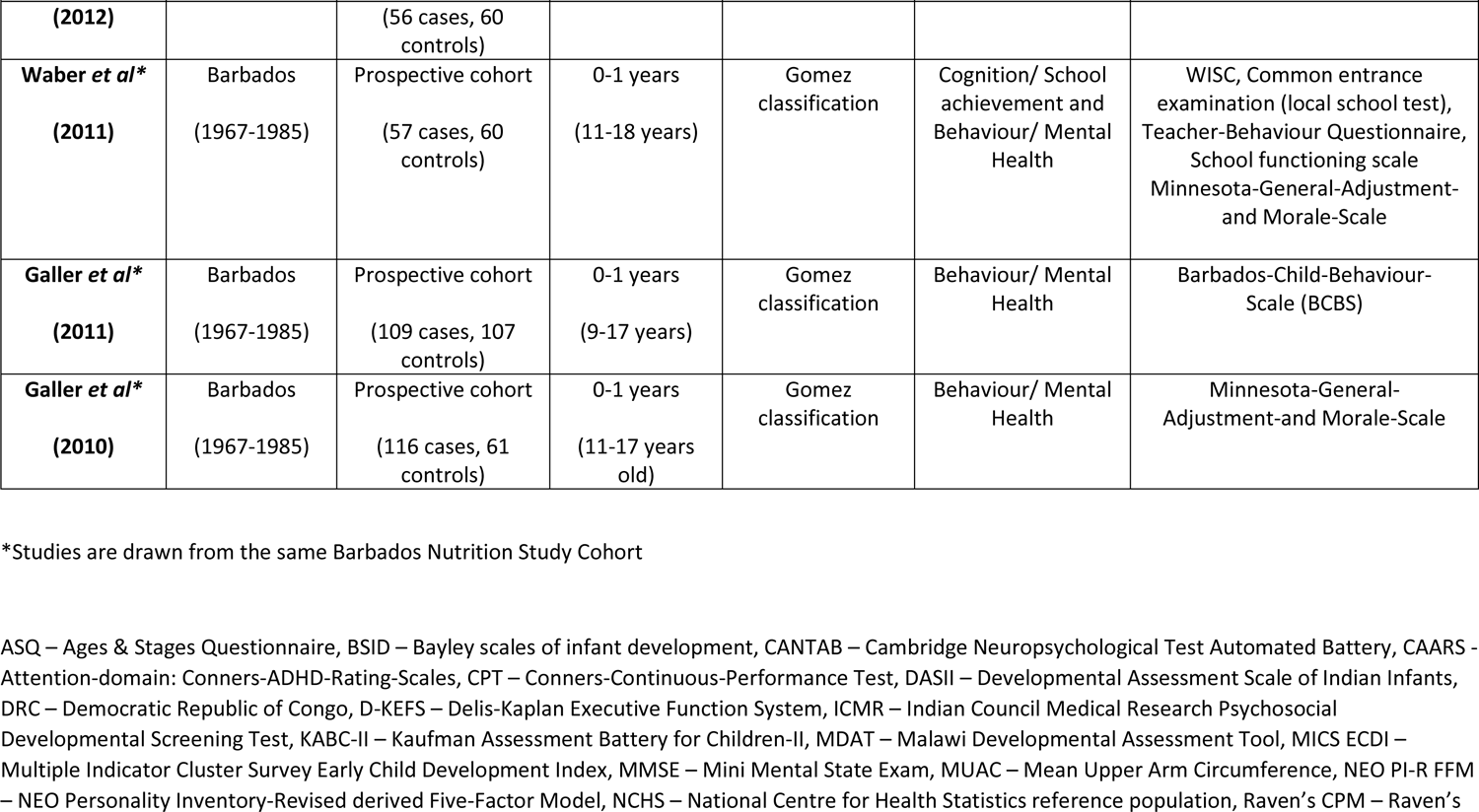
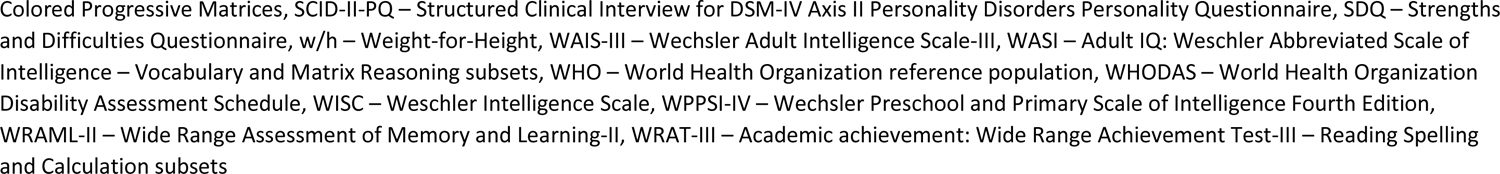
Study Characteristics

**Supplementary Table 2.**
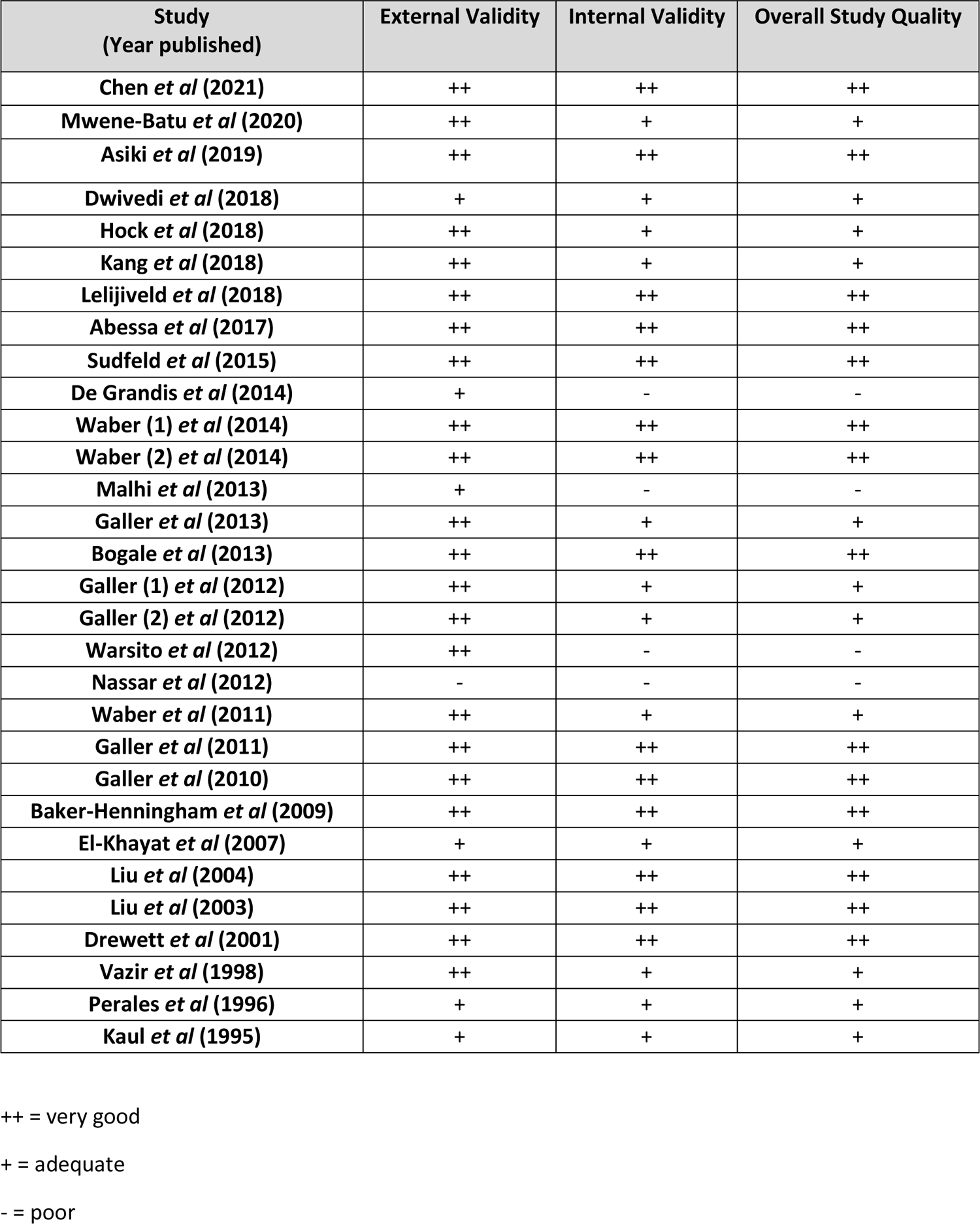
Study quality scoring

**Supplementary Table 3.**
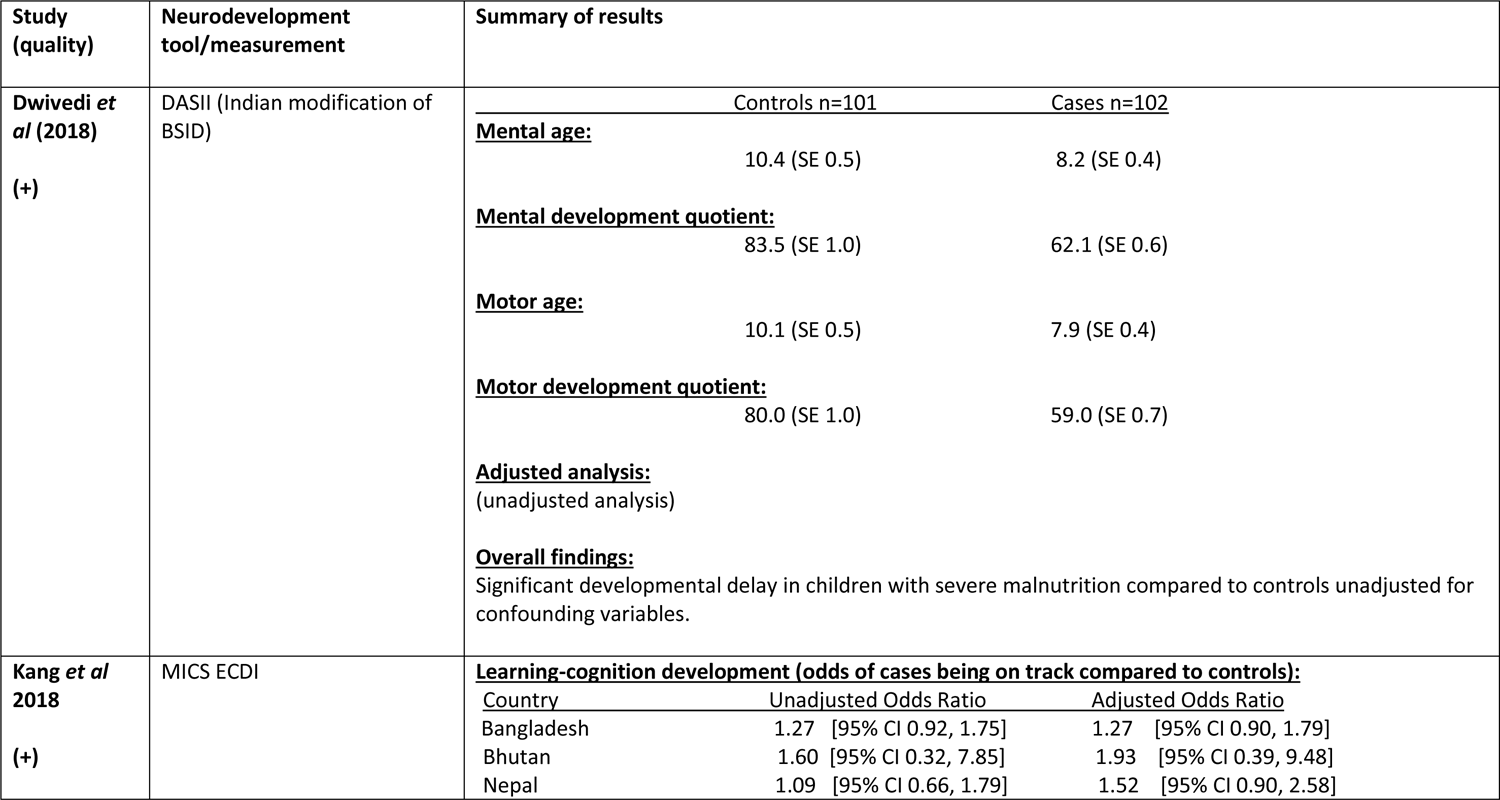
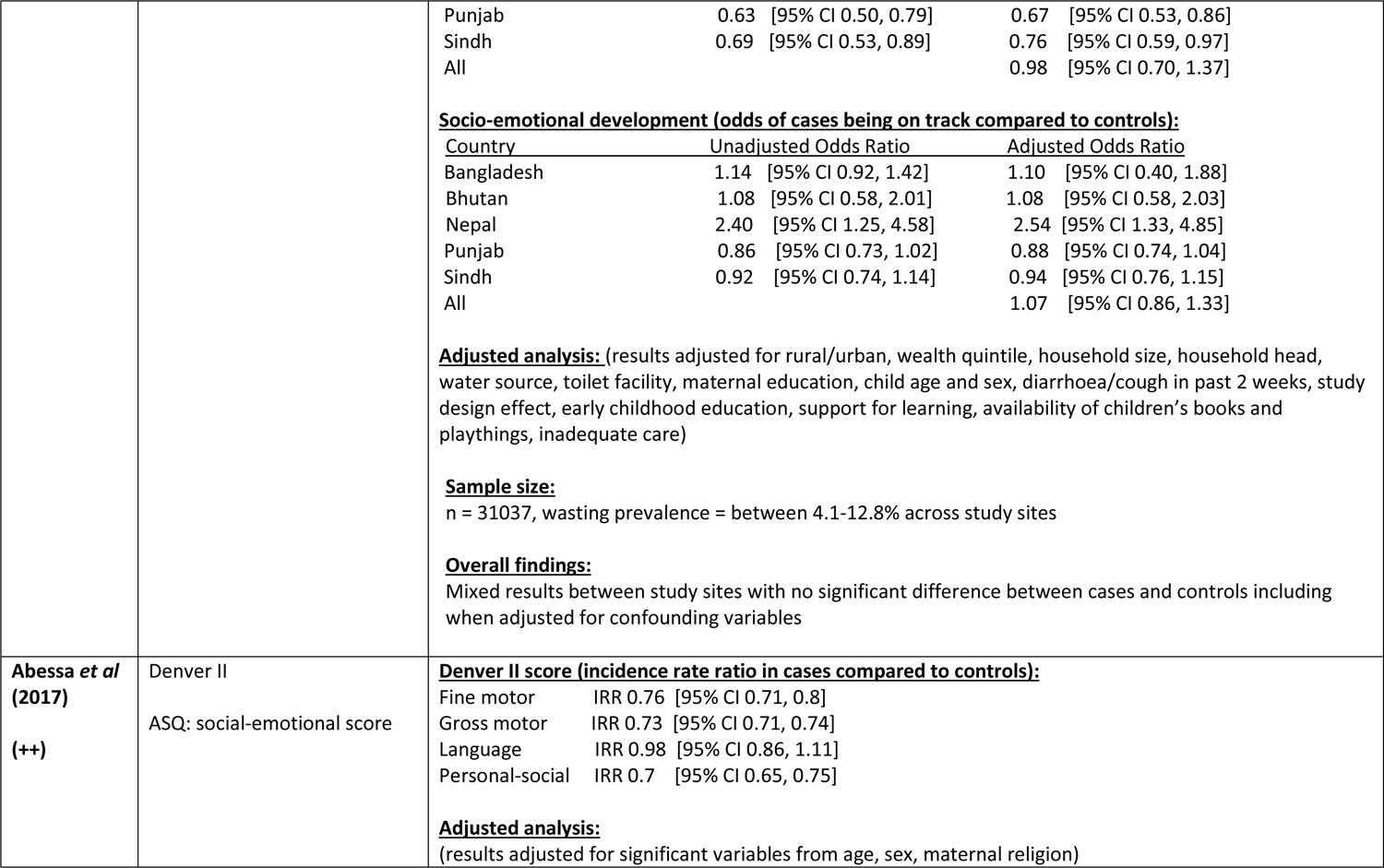
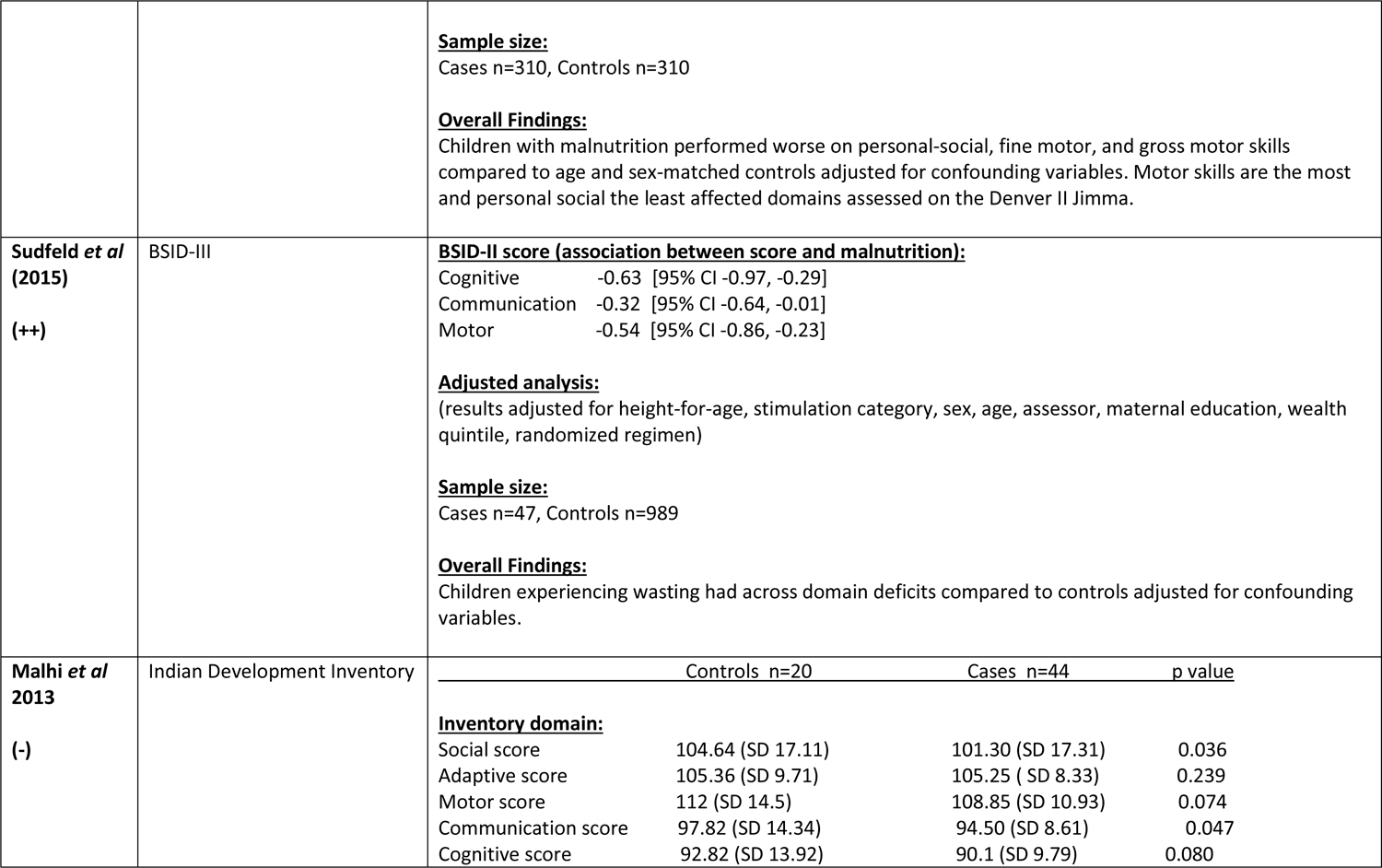
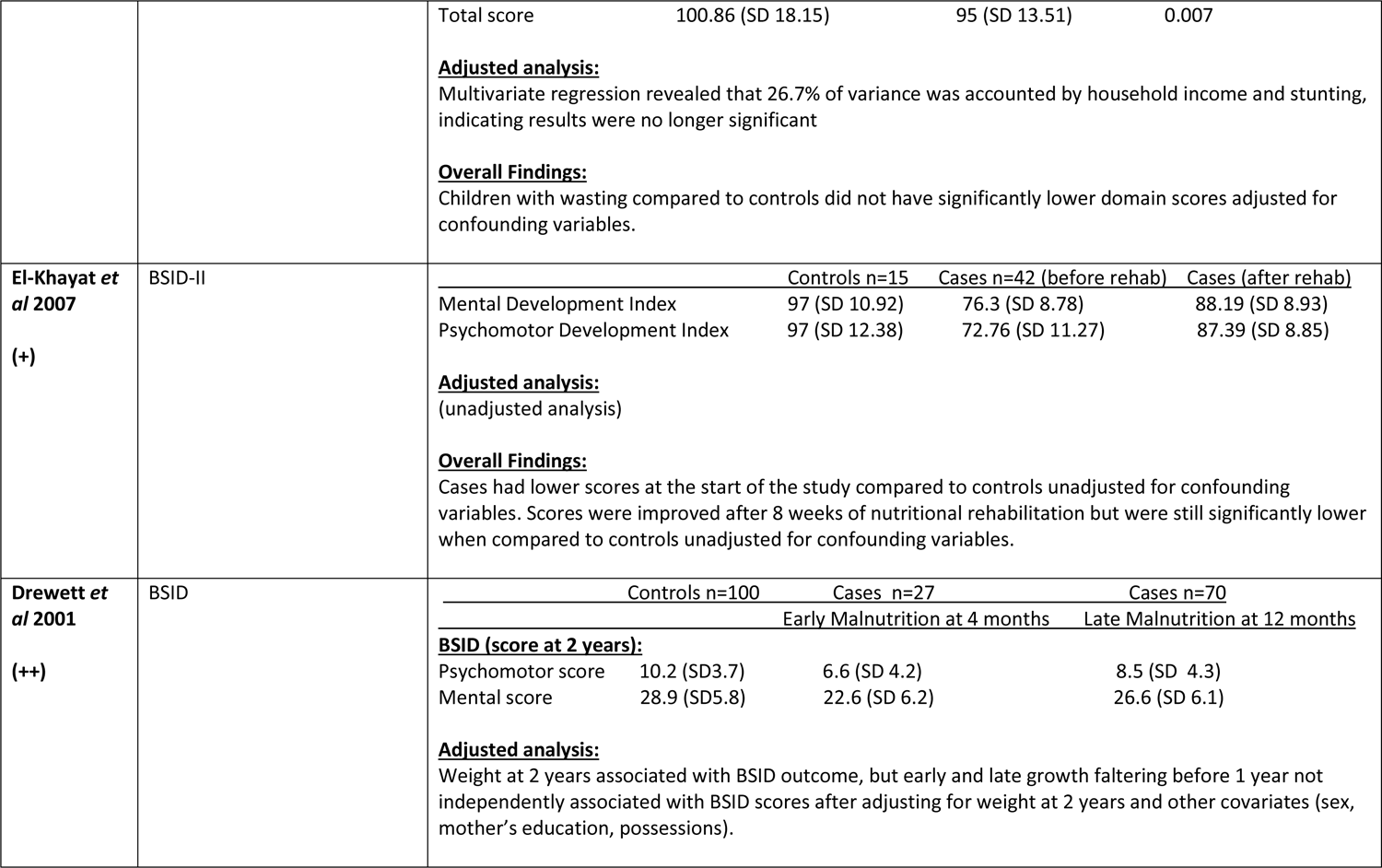
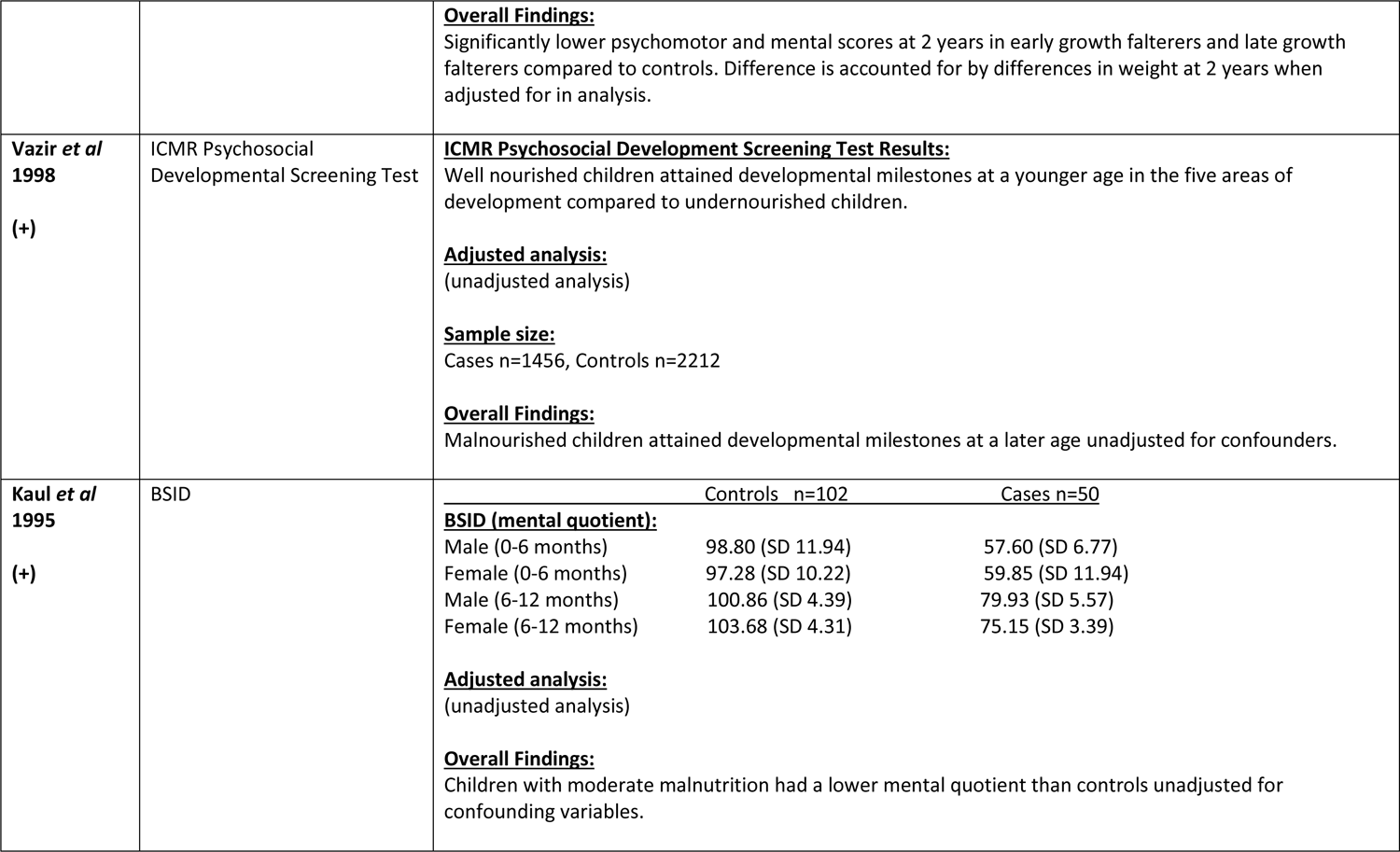
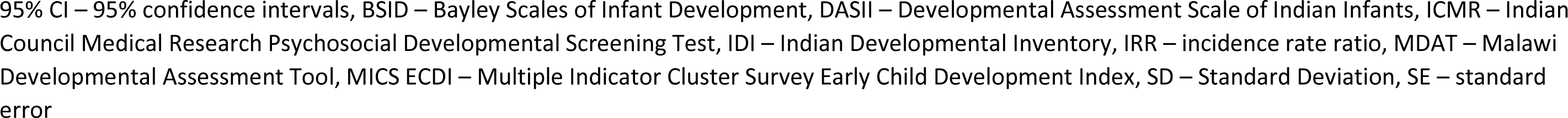
Results from studies assessing neurodevelopment in children with malnutrition compared to controls

**Supplementary Table 4.**
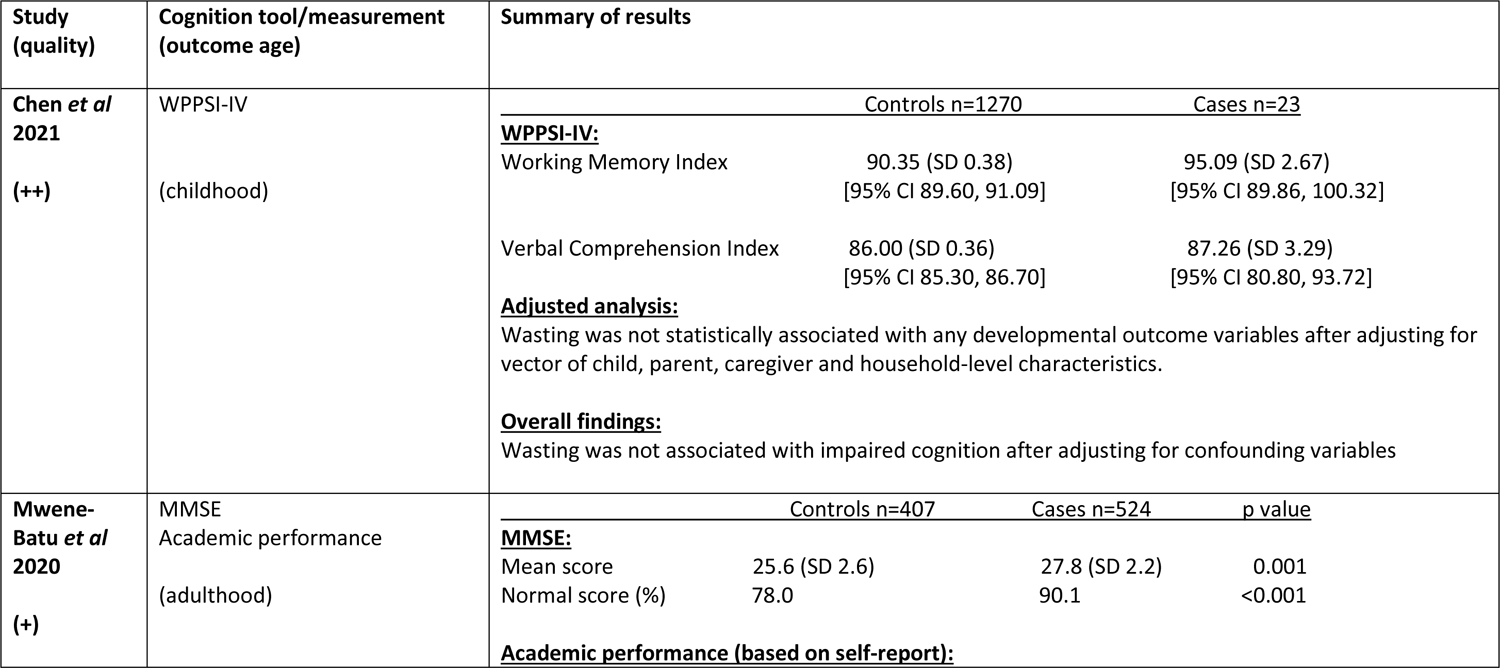
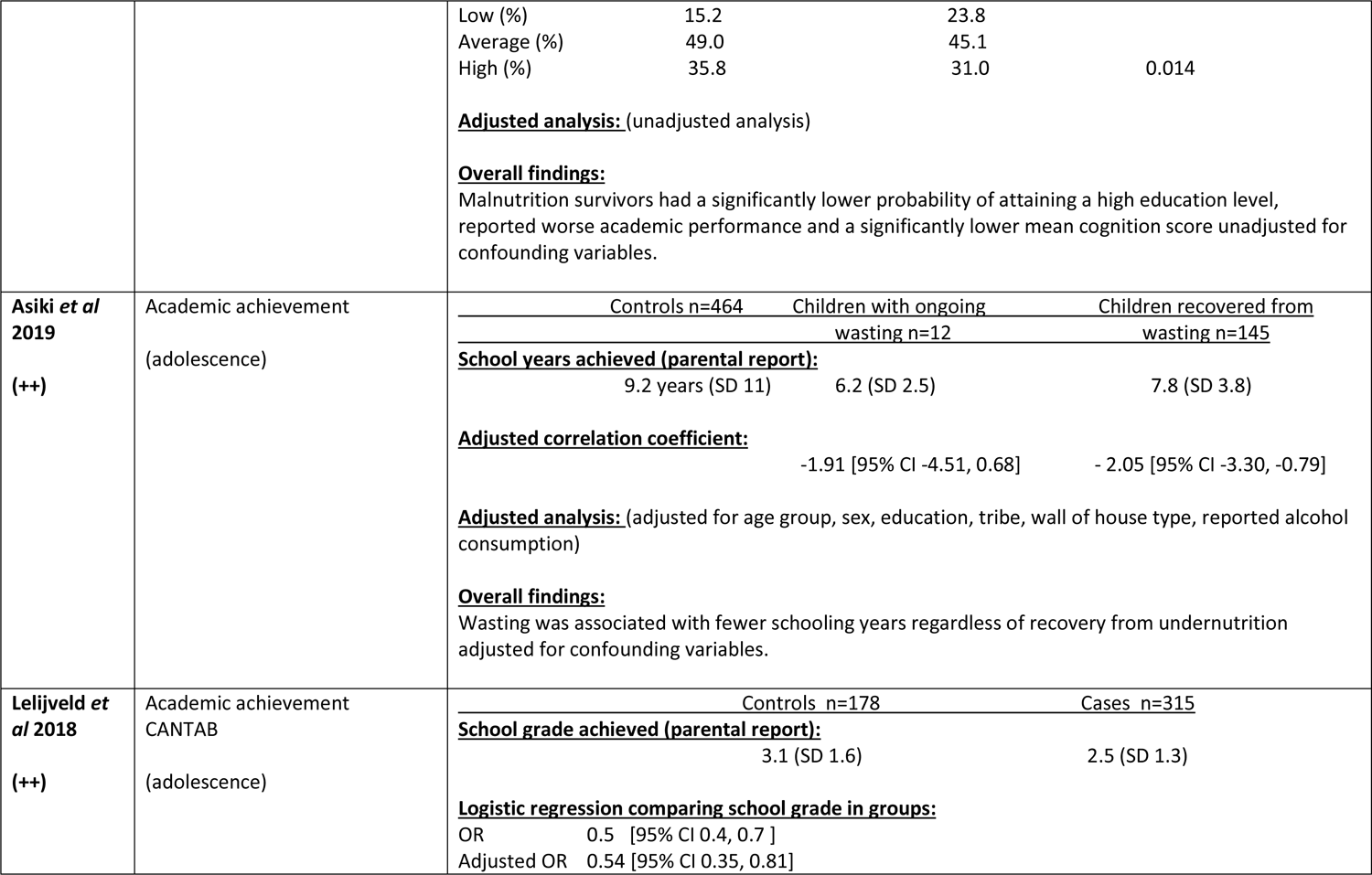
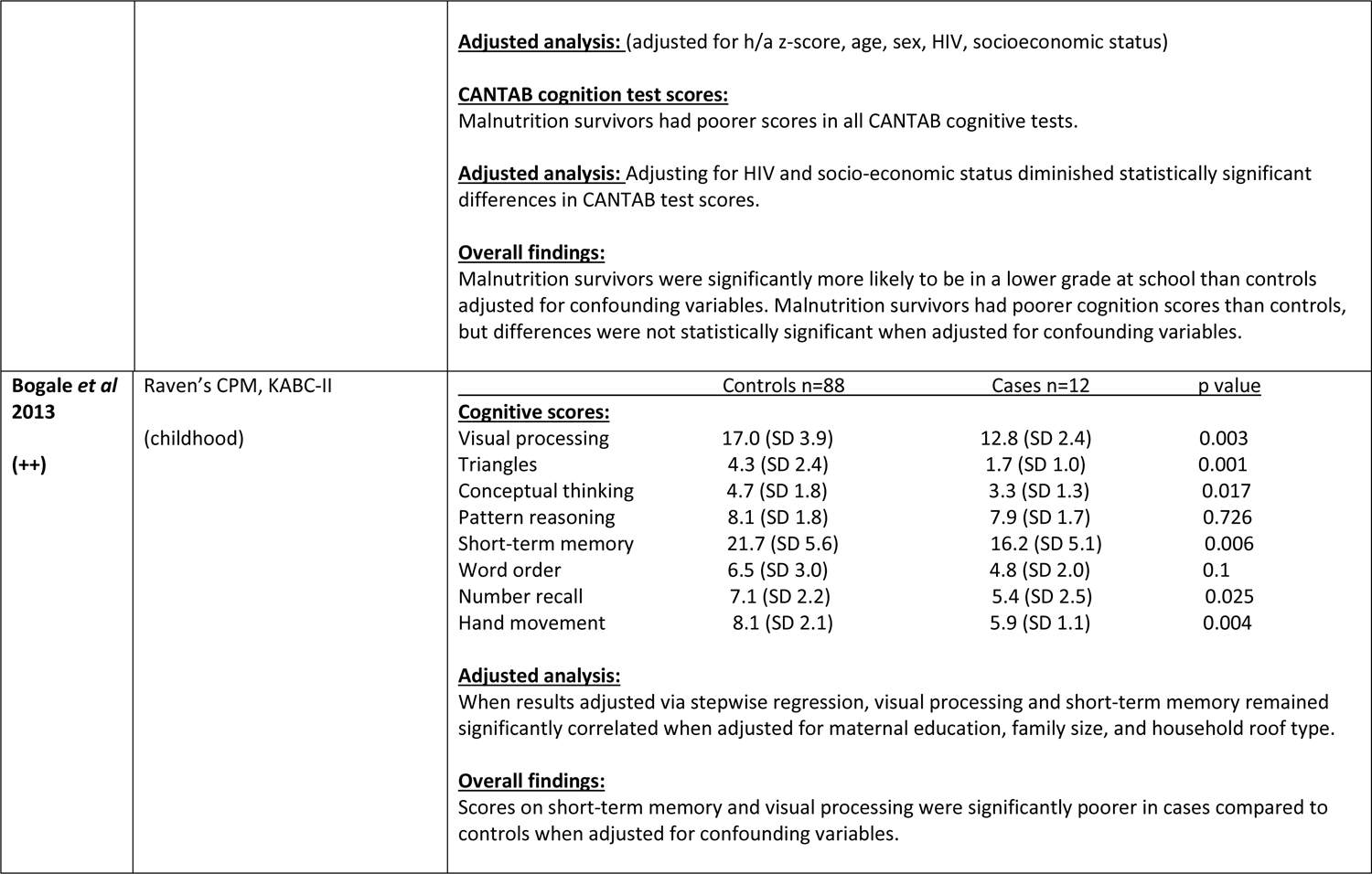
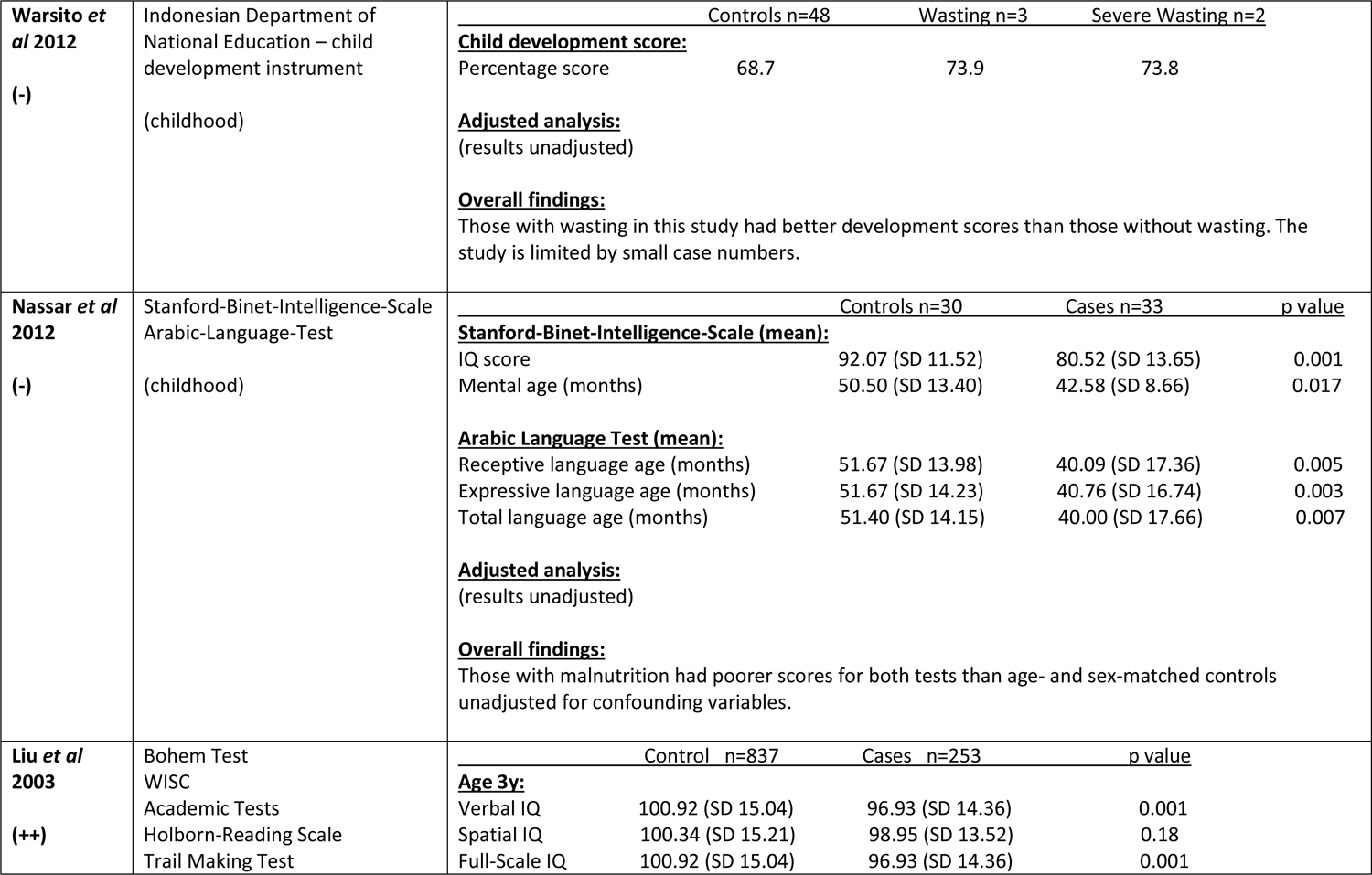
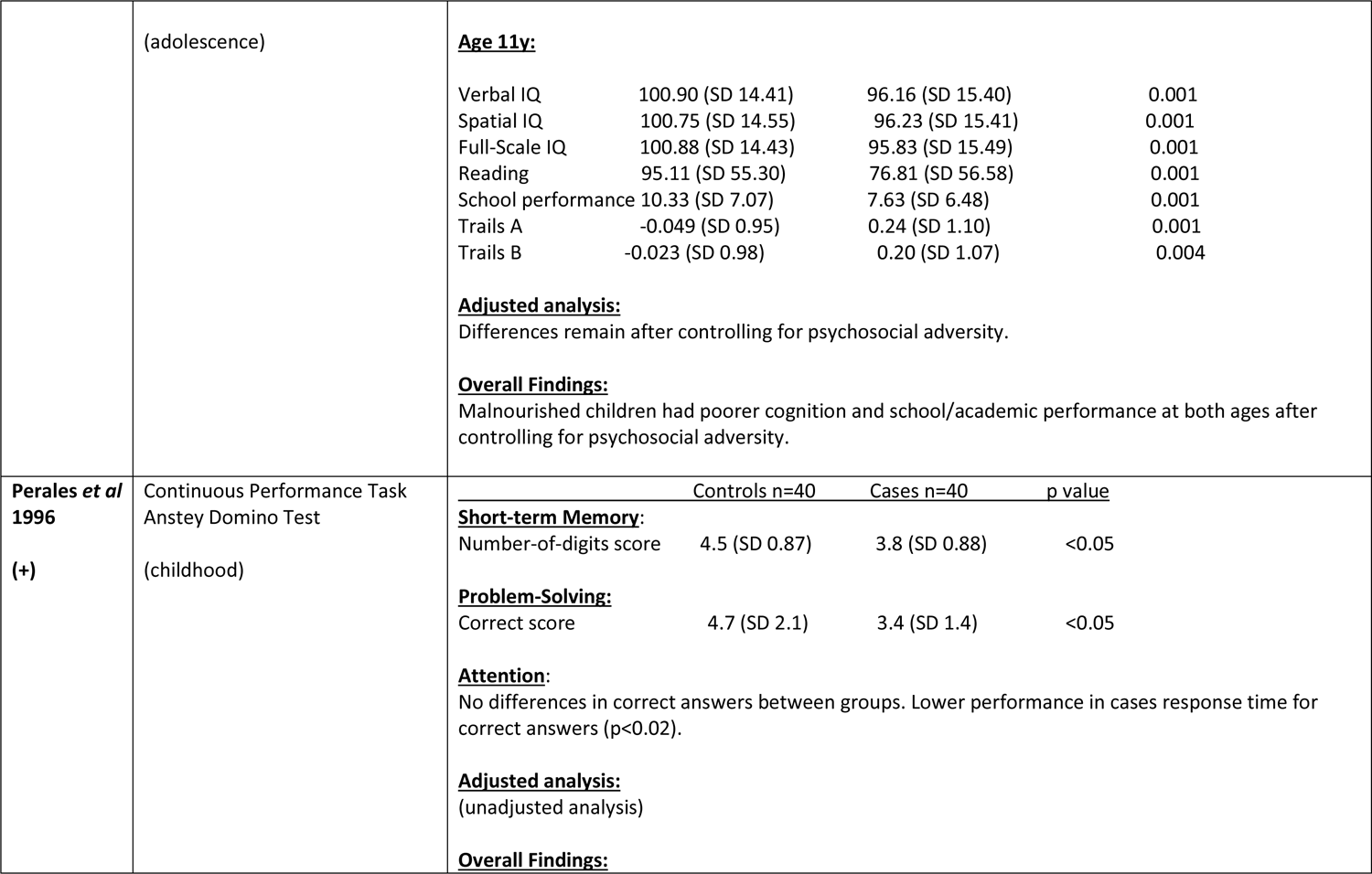
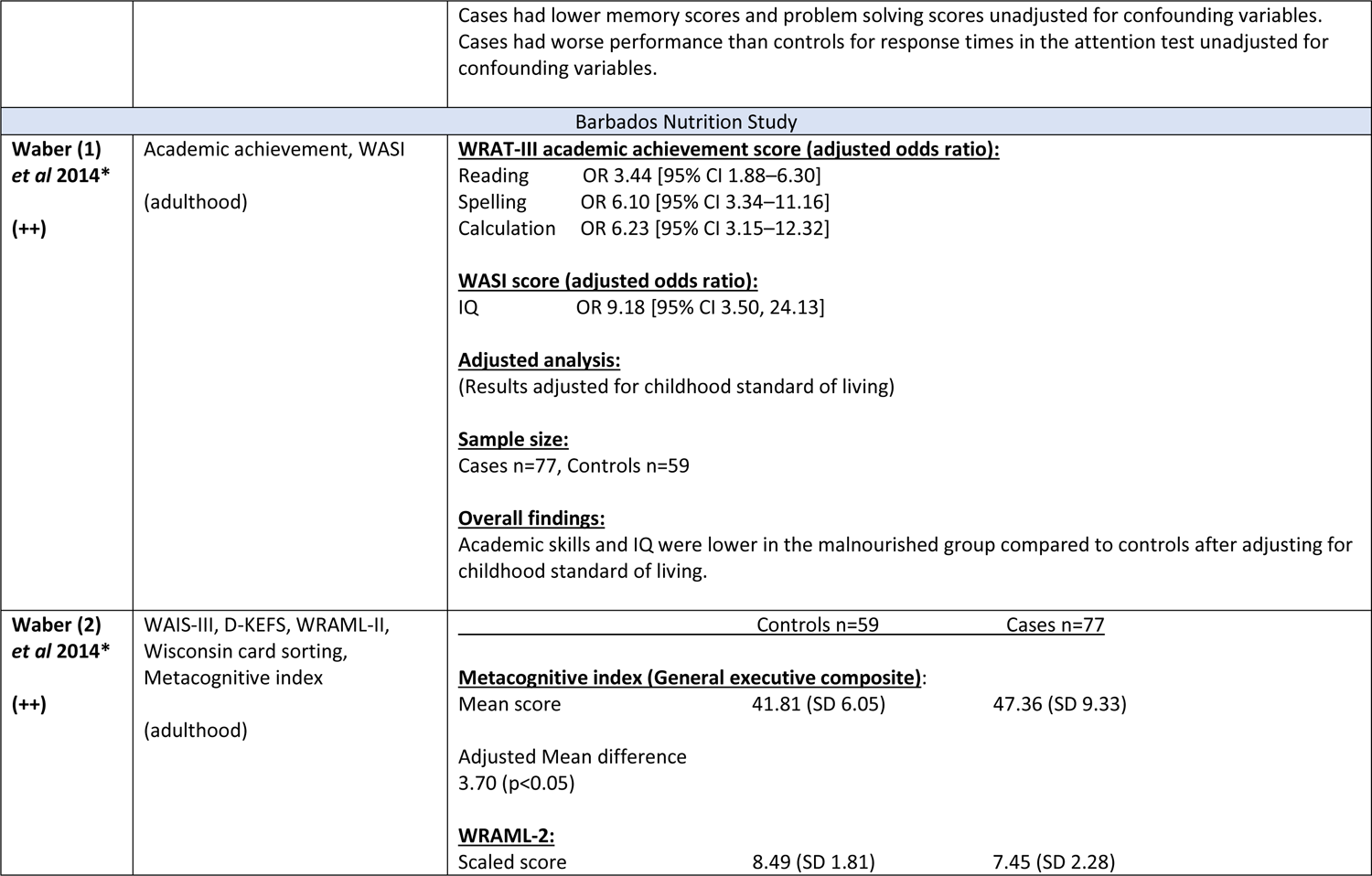
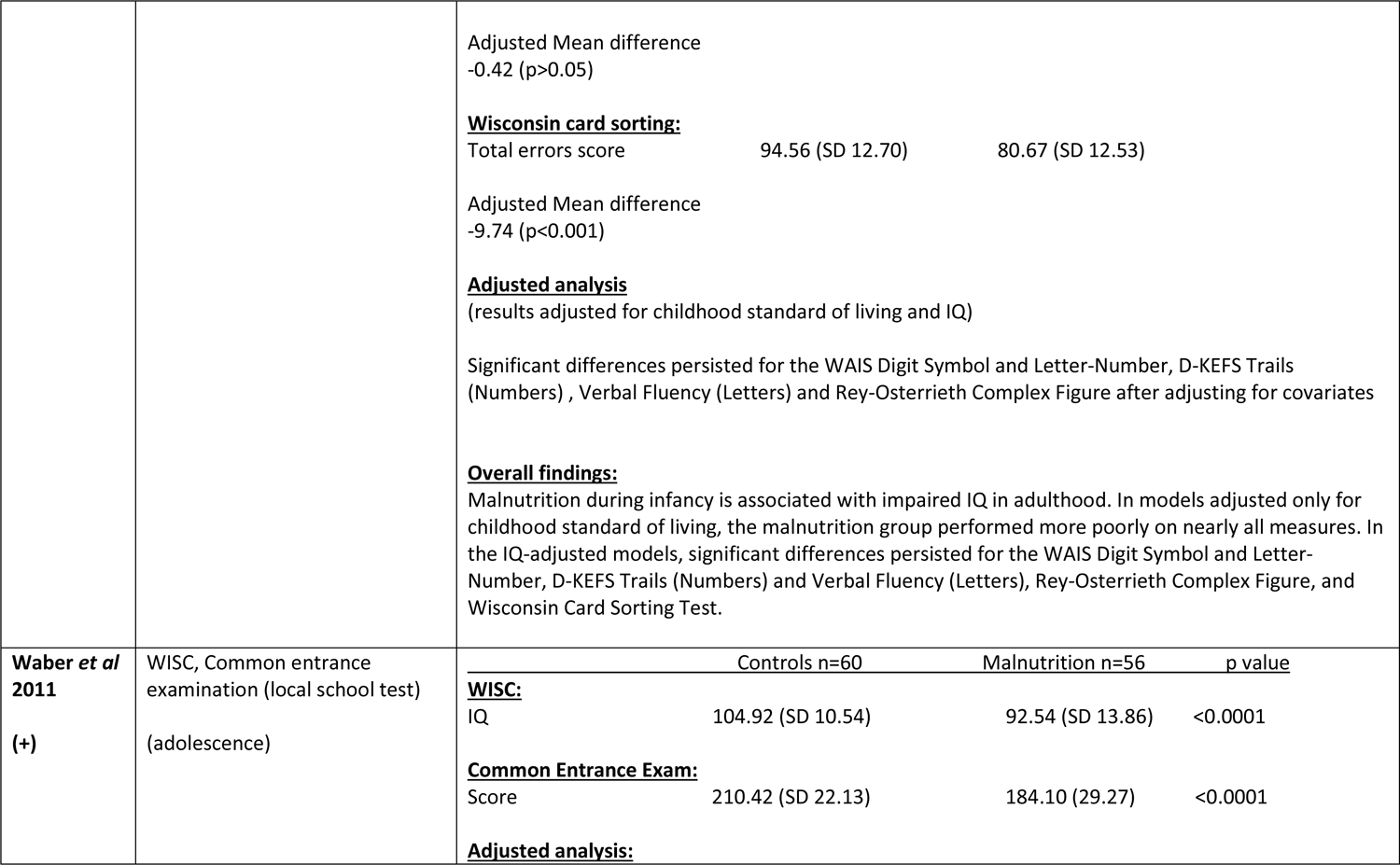
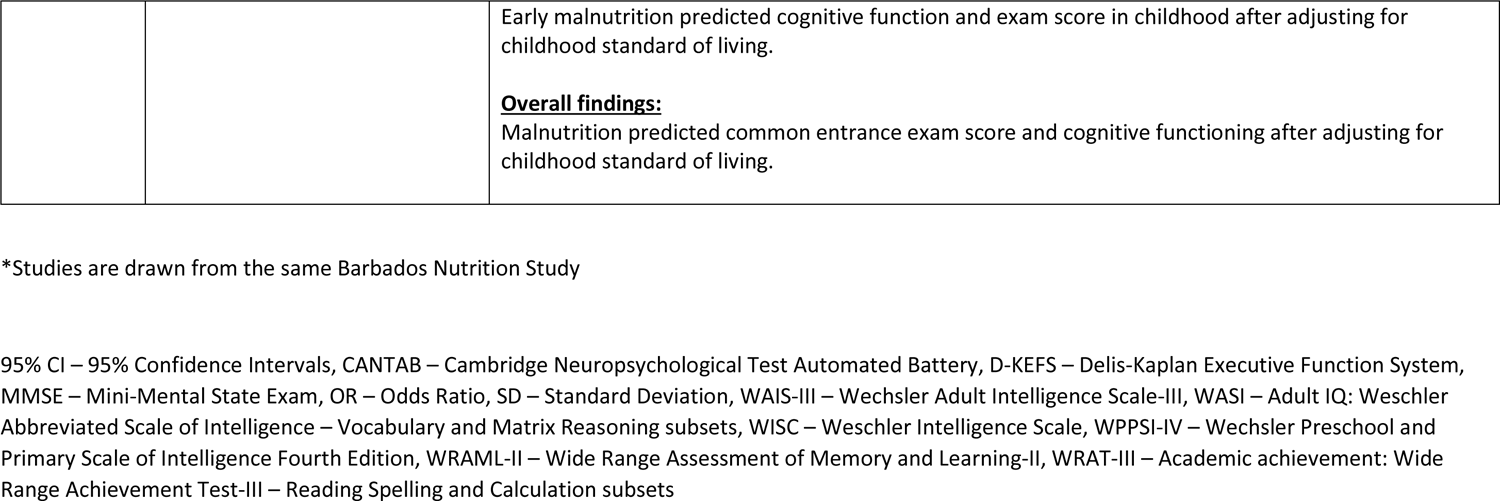
Results from studies assessing cognitive outcomes in those exposed to childhood malnutrition compared to controls

**Supplementary Table 5.**
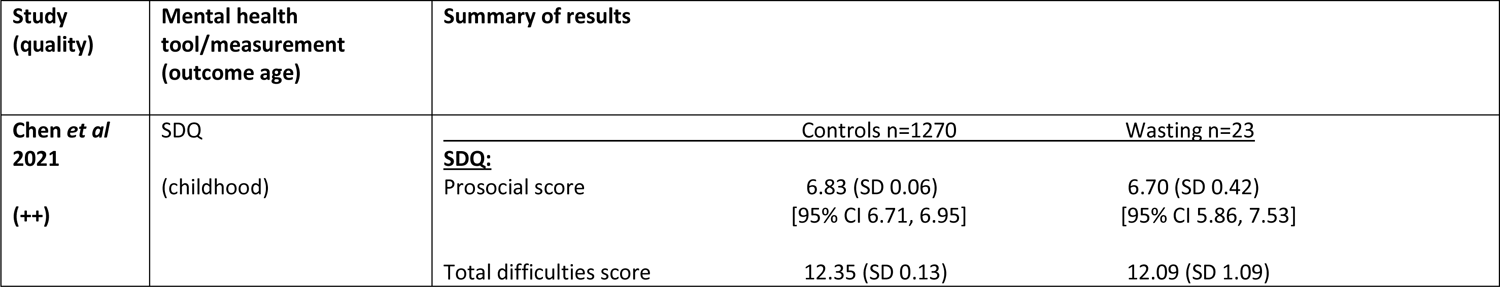
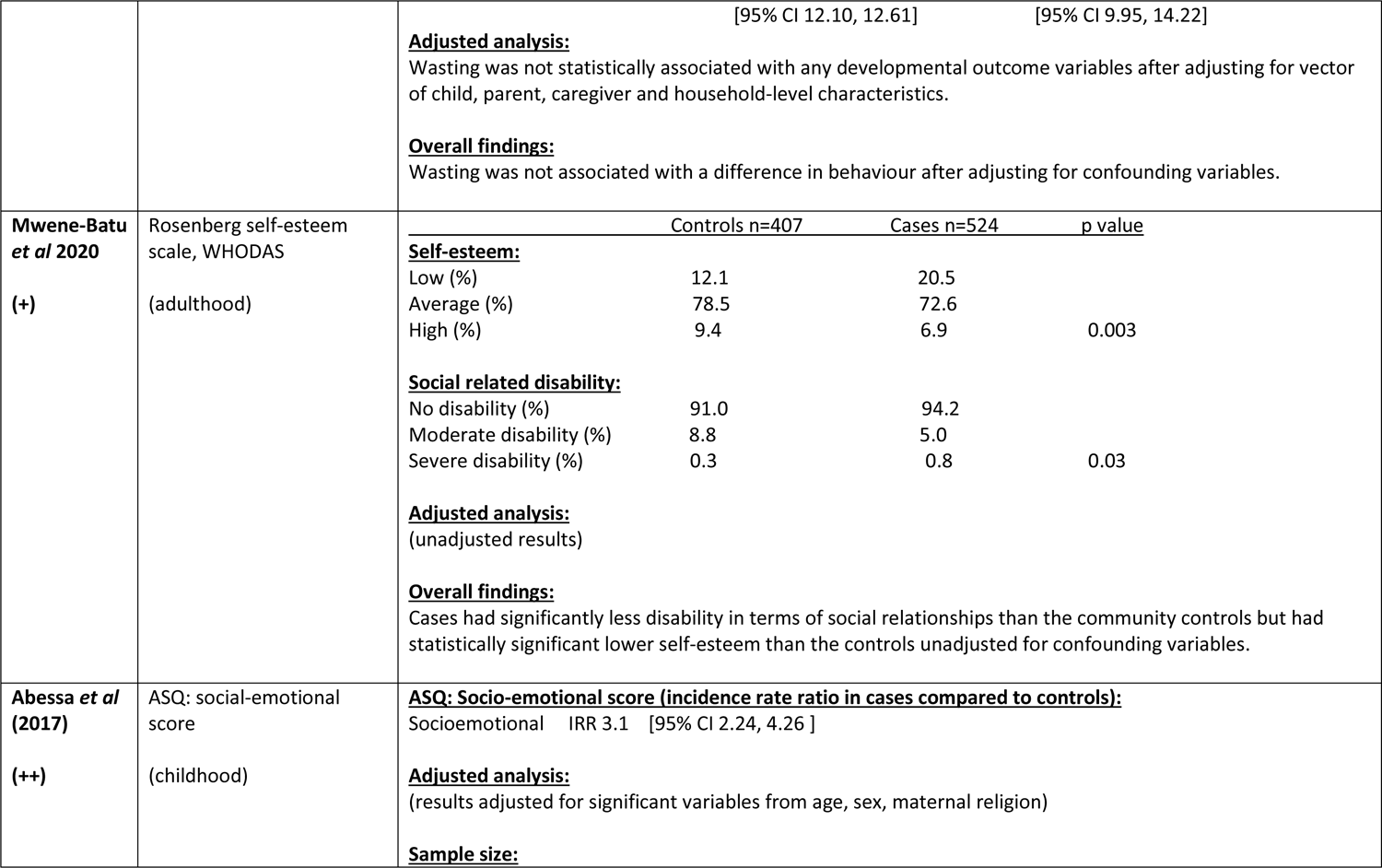
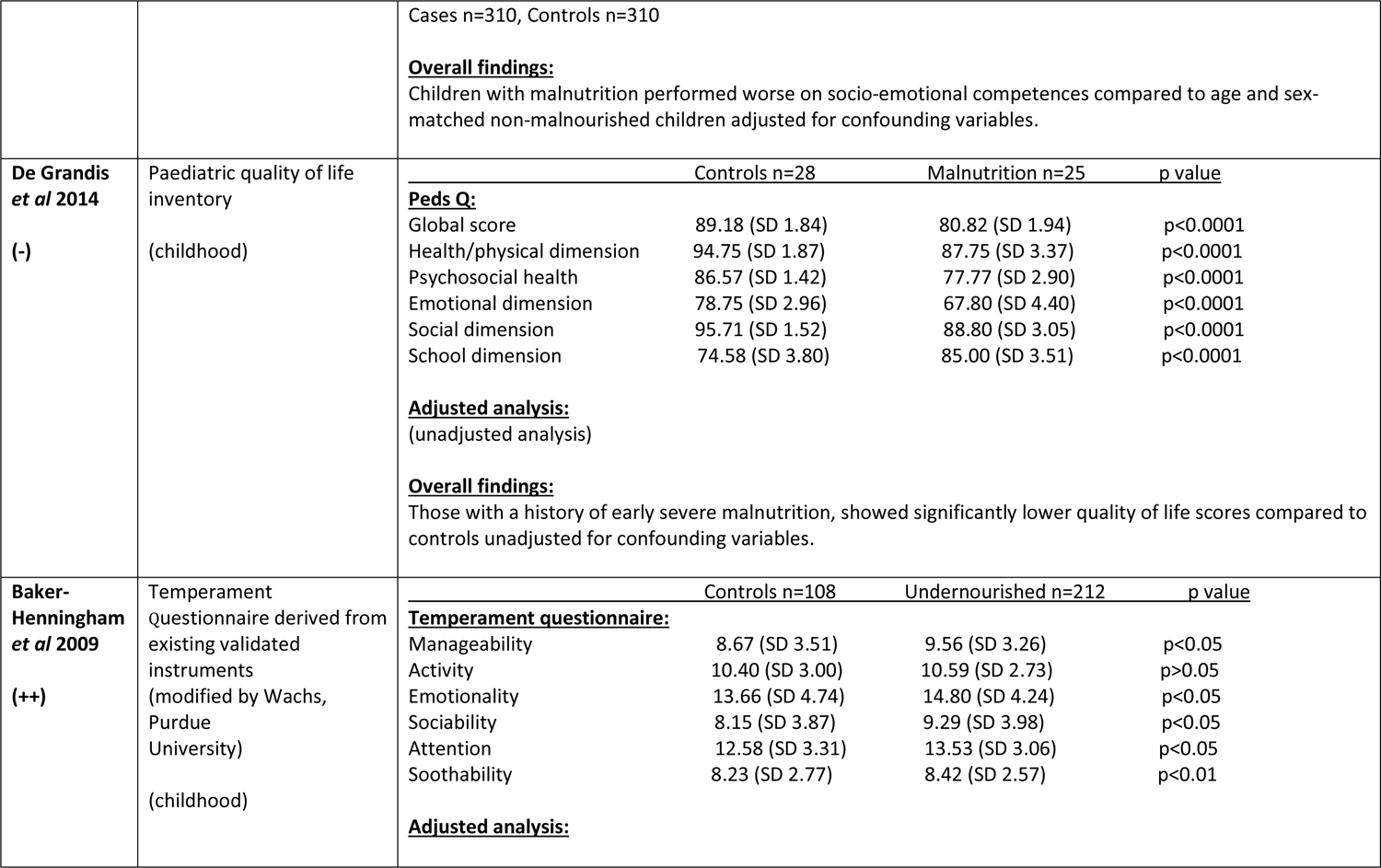
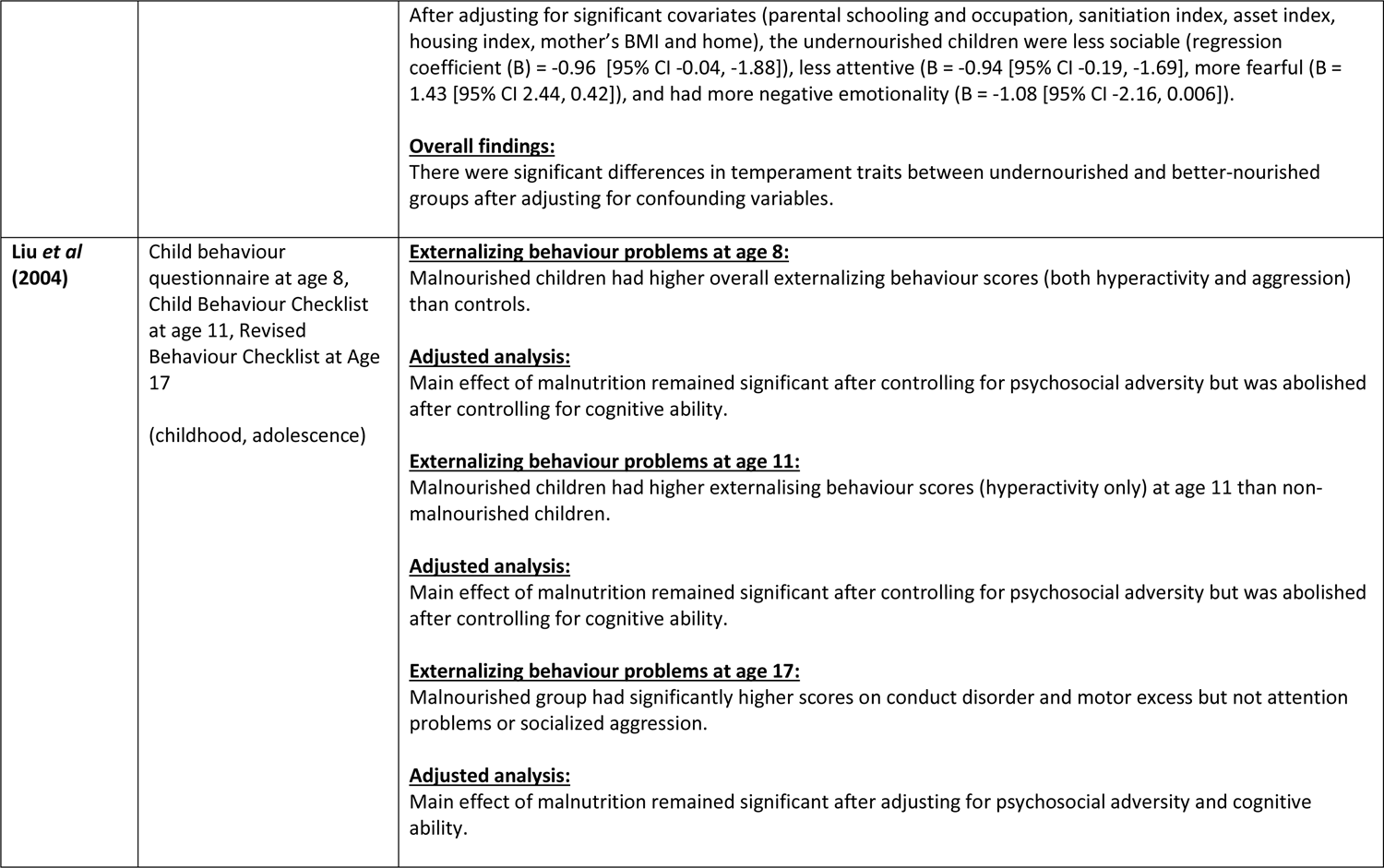
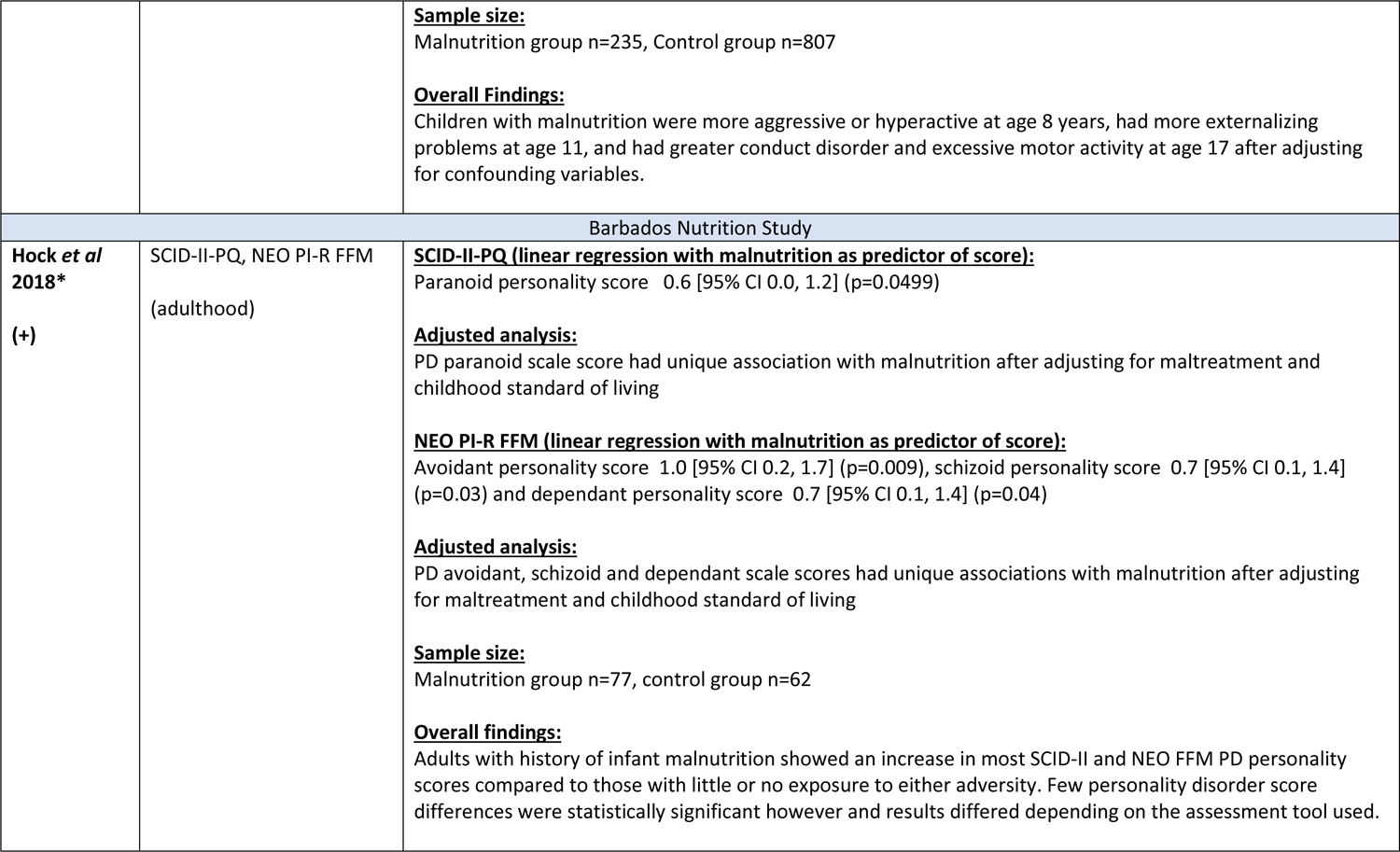
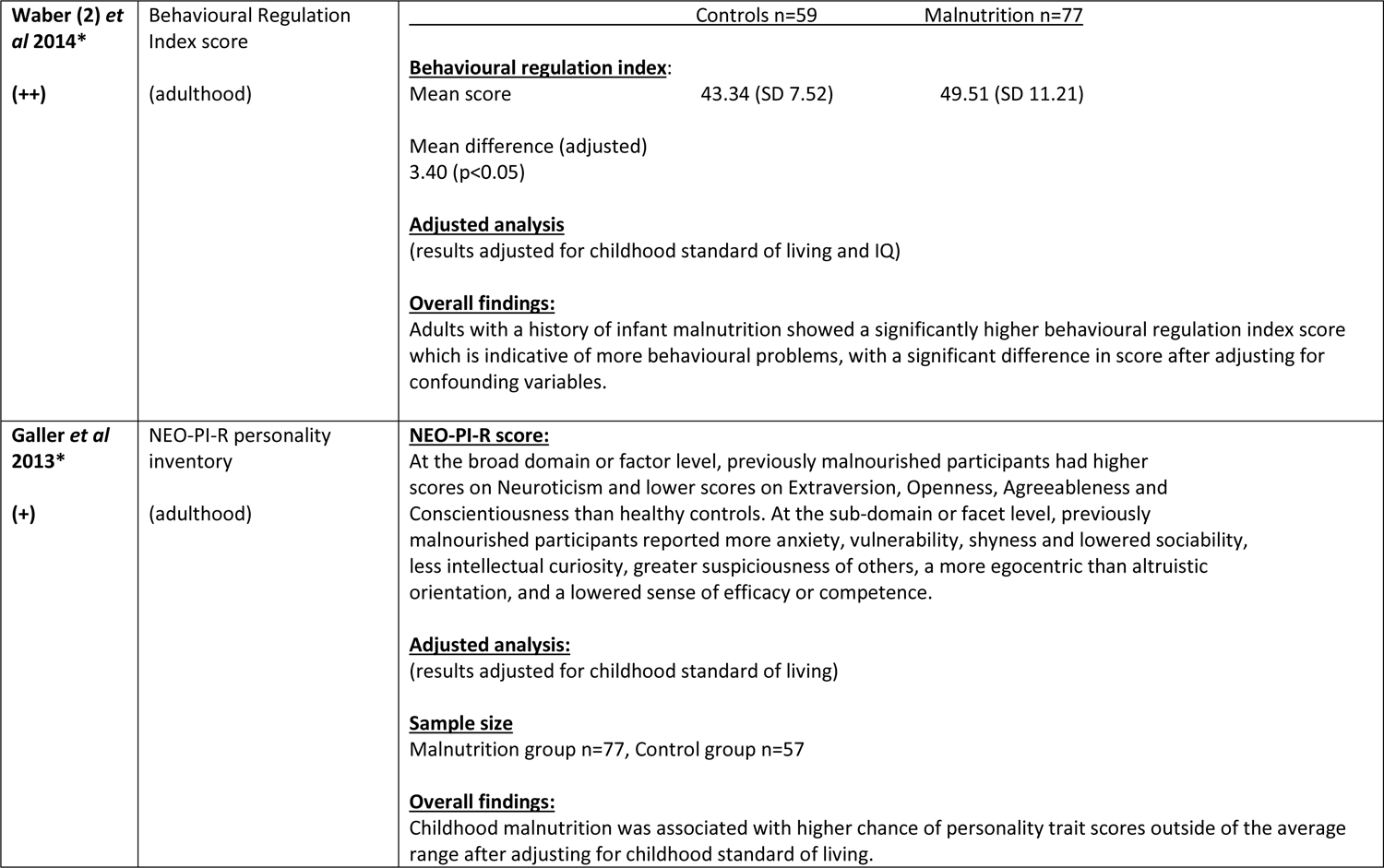
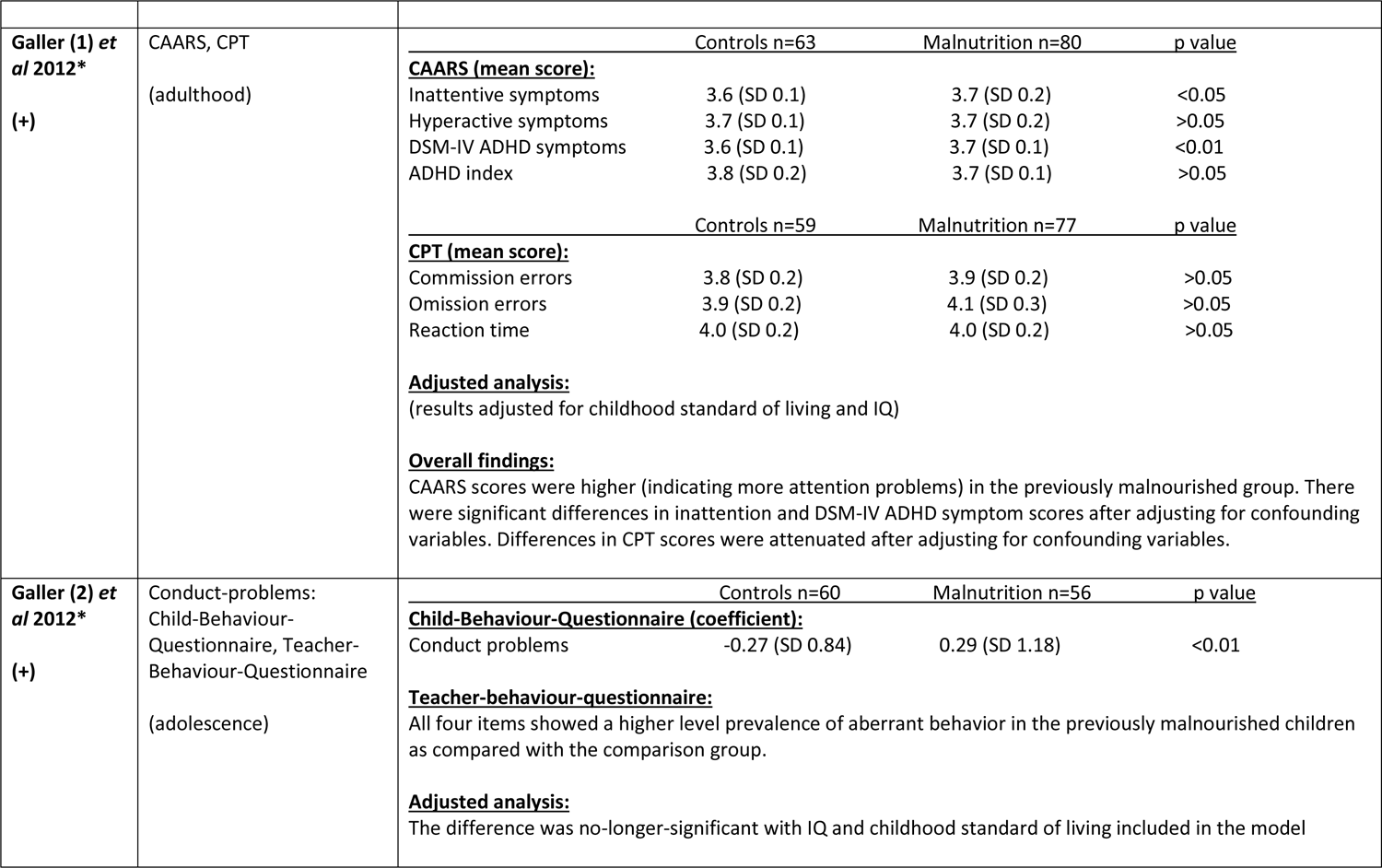
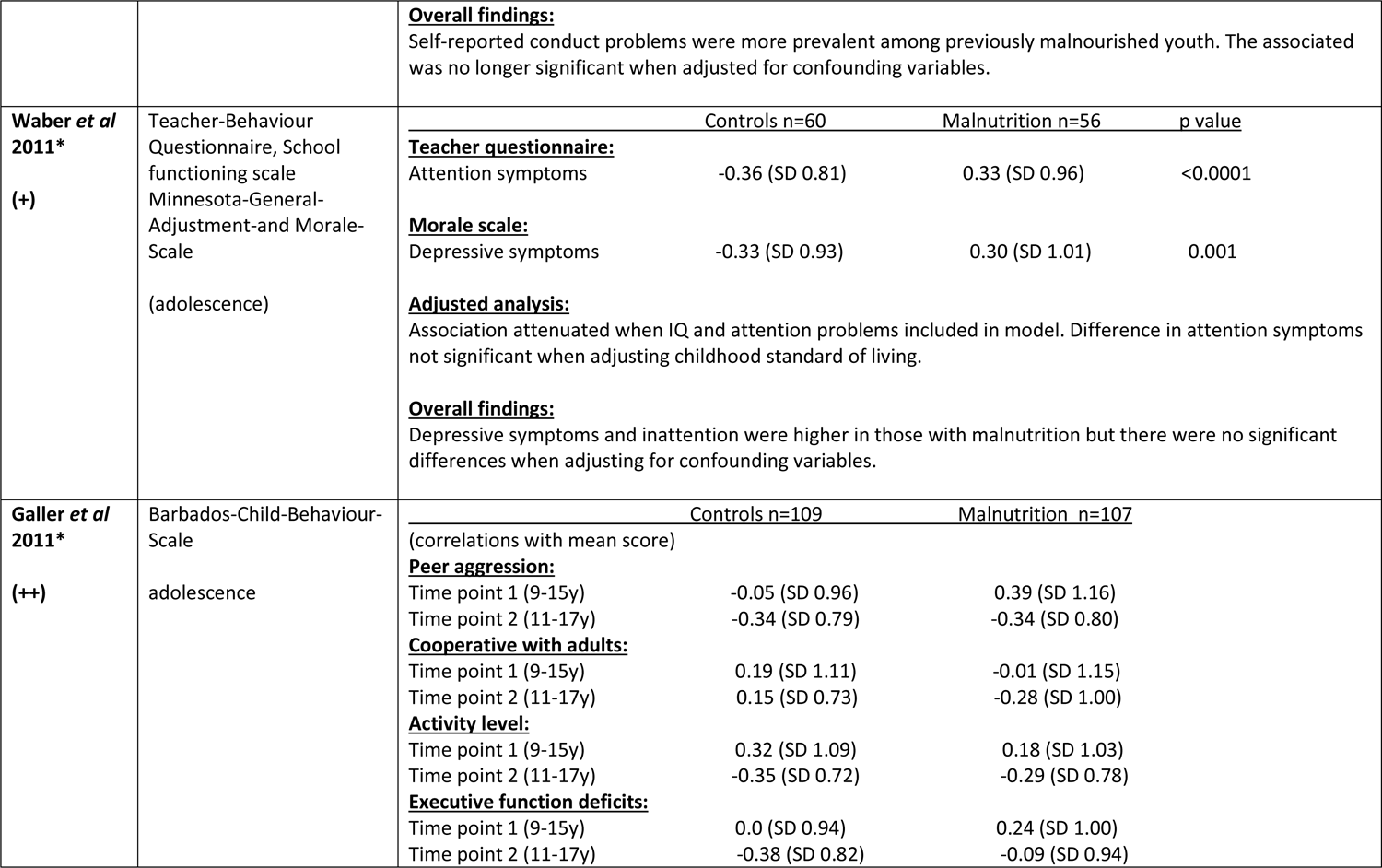
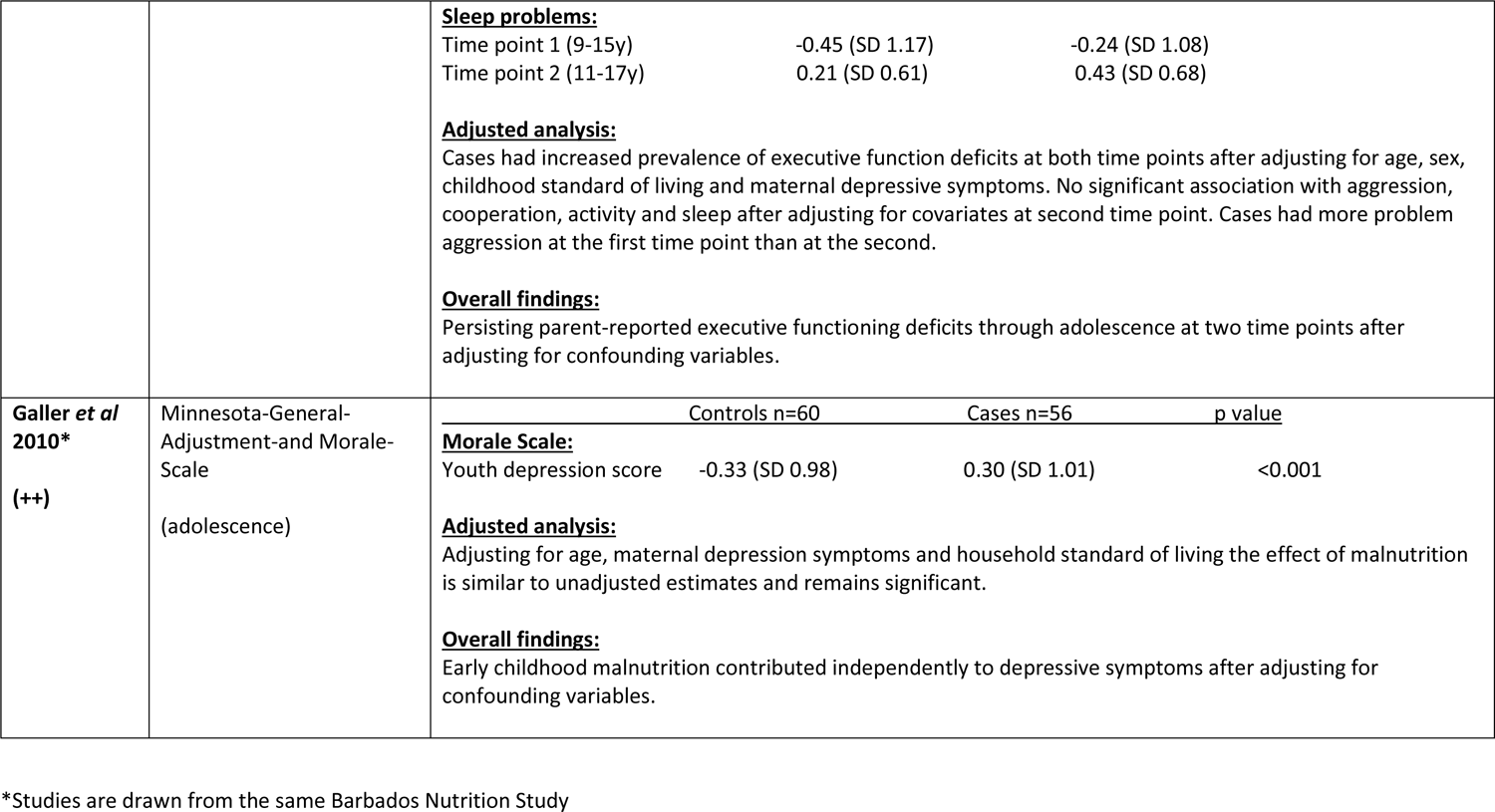
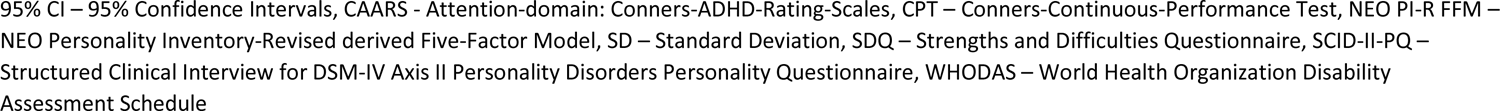
Results from studies assessing mental health outcomes in those exposed to childhood malnutrition compared to controls

